# Modeling COVID-19 dynamics in Illinois under non-pharmaceutical interventions

**DOI:** 10.1101/2020.06.03.20120691

**Authors:** George N. Wong, Zachary J. Weiner, Alexei V. Tkachenko, Ahmed Elbanna, Sergei Maslov, Nigel Goldenfeld

**Affiliations:** Department of Physics, University of Illinois at Urbana-Champaign, Urbana, IL 61801, USA; Center for Functional Nanomaterials, Brookhaven National Laboratory, Upton, NY 11973, USA; Department of Civil Engineering, University of Illinois at Urbana-Champaign, Urbana, IL 61801, USA; Department of Bioengineering, University of Illinois at Urbana-Champaign, Urbana, IL 61801, USA; Carl R. Woese Institute for Genomic Biology, University of Illinois at Urbana-Champaign, Urbana, IL 61801, USA

## Abstract

We present modeling of the COVID-19 epidemic in Illinois, USA, capturing the implementation of a Stay-at-Home order and scenarios for its eventual release. We use a non-Markovian age-of-infection model that is capable of handling long and variable time delays without changing its model topology. Bayesian estimation of model parameters is carried out using Markov Chain Monte Carlo (MCMC) methods. This framework allows us to treat all available input information, including both the previously published parameters of the epidemic and available local data, in a uniform manner. To accurately model deaths as well as demand on the healthcare system, we calibrate our predictions to total and in-hospital deaths as well as hospital and ICU bed occupancy by COVID-19 patients. We apply this model not only to the state as a whole but also its sub-regions in order to account for the wide disparities in population size and density. Without prior information on non-pharmaceutical interventions (NPIs), the model independently reproduces a mitigation trend closely matching mobility data reported by Google and Unacast. Forward predictions of the model provide robust estimates of the peak position and severity and also enable forecasting the regional-dependent results of releasing Stay-at-Home orders. The resulting highly constrained narrative of the epidemic is able to provide estimates of its unseen progression and inform scenarios for sustainable monitoring and control of the epidemic.

---

On January 24, 2020, the second known COVID-19 case to be diagnosed in the USA was reported in Chicago, Illinois. Community transmission of the disease was confirmed on March 8, 2020. During the subsequent ten days, the epidemic grew with a case doubling time of approximately 2.3 days, while testing capacity was essentially fixed. On March 21, 2020, a Stay-at-Home order was issued for the entire state of Illinois and subsequently extended on March 31, 2020 and again on April 23, 2020. The order was lifted on May 30, 2020 [1]. Responsible relaxation of the mitigation of COVID-19 must be informed by realistic and well-calibrated epidemiological modeling of the outcomes of any scenarios under consideration—not just of the resulting (increased) death toll but also of the stress placed upon the healthcare system. The purpose of this report is to present such an analysis.

A variety of modeling approaches are used by hospitals, public health officials, and state governments. These range between phenomenological models that use a curve-fitting procedure to match data, such as the daily death rate, and mechanistic methods that model the trajectory of the epidemic as individuals transition through several disease and healthcare-bound stages [2-5]. Only mechanistic models are able to make justifiable predictions while accounting for changes in the epidemic environment, such as the imposition or relaxation of community mitigation efforts. Of these, compartmental models like the Susceptible-Infectious-Recovered (SIR) models, and Susceptible-Exposed-Infectious-Recovered (SEIR) extensions, are widely used. Compartmental models describe how fractions of a homogeneous, well-mixed population progress through different states the disease, driven by interactions between infectious and susceptible individuals. In the simplest models, the dynamics is deterministic and the rates are constant in time, but many variants and extensions exist and are widely used.

In order to be practically useful, models must be calibrated to observed data [4, 6-8]. We calibrate the important dynamics of the model to several simultaneous streams of empirical data including total and in-hospital deaths, as well as hospital and ICU bed occupancy by COVID-19 patients. To avoid biases resulting from nonuniform and non-constant testing rates, which may be difficult to parameterize, we do not consider positive case data. The resulting model is a description of the epidemic as it progresses through the hospital system in Illinois; as it is clear that a non-negligible number of COVID-19 deaths occur outside the hospital environment (e.g., in homes and nursing homes especially), we augment our model with an effective description of the net incidence of deaths due to COVID-19.

There are many limitations to the types of models that we and others use to describe COVID-19, and these have been explored extensively in the literature, especially with regard to spatial structure [9], superspreader events and individuals [10-13], and the structure of contact networks [14-16]. A geographical region as large as the state of Illinois is not well-described as homogeneous, due to large variations in population density and prevalence of infection between the Chicago metropolitan area and the more rural regions in Central and Southern Illinois. Indeed, most of the known cases and deaths to date have occurred in the Chicago area (by roughly by an order of magnitude), so in practice this region dominates our results for the state as a whole.

One cannot simply scale the results of models for the Chicago metropolitan area to the entire state, however, because the transmission characteristics depend on the frequency and duration of contacts, which likely vary significantly among geographical regions. For this reason, we simulate the dense urban areas and the three sparser, more rural areas separately. We note that a more refined treatment would account for transfers between these separate populations, as well as transfers into and out of the state as a whole; however, we do not currently model these processes. The number of cases in individual rural areas is sufficiently small, due to the early Stay-at-Home order, that a long phase of exponential growth in these regions was largely avoided. As a result, these populations are not well-described by continuum, deterministic models. Nevertheless, by aggregating these populations, the numbers are large enough that deterministic, exponential growth, at least at the early stages, was visible.

In this work, we describe an estimate of the rise and fall of the epidemic within Illinois, taking into account the modulation of the transmission parameters due to social distancing. In the following sections, we first describe our extension of the SEIR model, which takes into account the long and variable delay times reported in the literature. After describing the procedure by which we calibrate the model to data, we argue for the robustness of short term model predictions. Finally, we present and discuss our predictions of the epidemic trajectory through the Summer of 2020, including the effects of the release of the Stay-at-Home order at the end of May.

## I. MODEL DESCRIPTION

To facilitate a general treatment of delay times in the COVID-19 epidemic, we implement a non-Markovian model that derives from the classic Kermack-McKendrick age-of-infection model [17]. Age-of-infection models are similar to compartmental models, such as Susceptible-Infectious-Recovered (SIR) or Susceptible-Exposed-Infectious-Recovered (SEIR) models, but allow transition delays between entering sequential states to be drawn from arbitrary probability density functions (see, e.g. [4, 18]). We use non-Markovian models because their delays can be defined by an arbitrary number of timescales, in contrast to the single exponential rate parameter of compartment based models. Non-Markovian models are thus, in principle, better able to reproduce the observed signal delays between different states, e.g., the flattening of the curve of hospital admissions compared to in-hospital deaths.

The use of deterministic rather than stochastic epidemic descriptions is generally justified when the modeled populations are large enough that relative daily changes are small and the number of individuals is large relative to one. This means that a deterministic compartmental model has a self-consistency check, because once the epidemic size is of order unity, a stochastic model allows the epidemic to die out; in contrast, in a deterministic model an epidemic will continue to evolve even with a unrealistic, fractional number of infectious individuals. We will see that for some regions of Illinois, our estimates are at the limit of validity of deterministic models.

We adapt the conventional age-of-infection model to include a number of delayed healthcare system observables of the epidemic, such as the number of patients in hospitals, Intensive Care Units (ICUs), and ultimately, the number of deaths.

### A. Time-modulated age-of-infection model

The core of our model is the daily incidence (i.e., the number of newly-infected individuals) in demographic (age) group *i, j_i_*(*t*). This value determines the dynamics of susceptible individuals in that group *S_i_*(*t*) according to

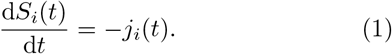

The incidence itself follows the renewal equation,

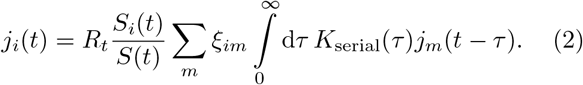

Here, *R_t_* is time-dependent effective reproduction number, 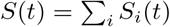 is the total susceptible population, *K*_serial_(*τ*) is the probability density function (PDF) of serial intervals, and *ξ_im_* is the contact matrix. For simplicity, we assume *ξ_im_* = 1, i.e., all demographic groups infect each other at the same rate. We assume that *N_i_*, the total number of individuals in the demographic group *i*, is approximately constant for the duration of the epidemic. In practice, this means our model neglects the effects of births, deaths due to causes unrelated to COVID-19, and mobility of the population, which is appropriate for the short time scales we model. We further assume that individuals are only infected once, i.e., that the duration of immunity to COVID-19 is longer than the timescale over which we simulate the epidemic. Our simulations begin on a day *t_s_* (whose value we sample during parameter inference) at which point we impose that ten people are spontaneously infected, setting *j_i_*(*t*) in proportion to the age distribution of the population under study.

We parameterize the effective reproduction number *R_t_* in terms of the basic reproduction number *R*_0_, a seasonal forcing factor *F*(*t*), a mitigation factor *M*(*t*), and the susceptible population fraction *S*(*t*)*/N* according to

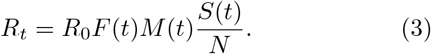

The formal dependence of *R_t_* on the total susceptible population *S*(*t*) is corrected in Eq. 2 by a factor *S_i_*(*t*)*/S*(*t*) which accounts for possible variation of susceptibility between different demographic groups. In our model, the homogeneous factor *M*(*t*) accounts for all sources of mitigation, including the effects of self-imposed isolation as well as government-mandated Stay-at-Home (SAH) orders, school closures, etc. We parameterize *M*(*t*) as a piecewise cubic Hermite interpolating polynomial that smoothly interpolates from 1 at *t*_0_ to *M*(*t*_1_) at *t*_1_ and is otherwise constant. The mitigation factor *M*(*t*_1_), start time, and end time of the interpolation interval are fitted by our algorithm. We choose to parameterize *M*(*t*) as a single transition (i.e., a single event) since this minimal model reduces the risk of overfitting to spurious trends. This choice is supported by the following observations: the adoption of social distancing practices took place relatively rapidly over a one to two week period; and the Stay-at-Home order remained active after the initial transition, presumably suppressing the magnitude of changes to mitigation. We will observe that this choice is a sufficient approximation for the duration of the data we use for calibration. We evaluate these claims more explicitly Section III D.

To model seasonal effects, we follow Ref. [19], which estimates seasonal forcing from the observed seasonal variability of positive tests in three other endemic coronaviruses. We thus adopt the functional form

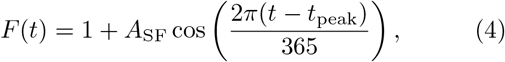

where *A*_SF_ denotes the strength of the forcing and *t*_peak_ sets the day of the year when seasonal forcing is strongest. From Ref. [19], we infer that seasonal forcing is strongest in the winter and set *t*_peak_ = January 16.

Ref. [19] finds evidence for *A*_SF_ = 0.2; however, we account for uncertainty in this parameter by sampling over *A*_SF_ during parameter inference. Incorporating this uncertainty is critical: if mitigation were only able to reduce the effective reproduction number to roughly unity, then seasonal forcing could drive a second wave of the epidemic.

For timescales of only a few months, our parameterization of *R_t_* includes a degeneracy: a change in the parameter *A*_SF_ may be compensated by adjusting the mitigation profile *M*(*t*). The degeneracy is broken as data is collected over long timescales, since *M*(*t*) models relatively instantaneous changes in infectiousness and *F*(*t*) produces an explicitly year-long, periodic modulation. Practically, this implies that *A*_SF_ may account for both seasonal effects and concurrent slow variations in the mitigation factor.

### B. Model topology

Due to limited and biased testing, neither the susceptible population *S_i_*(*t*) nor the daily incidence of new infection cases *j_i_*(*t*) are directly observable. Hence, we are forced to infer the dynamics of the epidemic using lagging and indirect indicators. These indicators include the total number of hospitalized (but not critical) patients *H*(*t*), the number of critically ill patients currently in ICU beds *C*(*t*), and *D*(*t*), the cumulative number of daily deaths in the hospital.

Our model topology assumes that all hospital deaths occur in ICU rooms. In practice, this simplification would be invalid if either a significant number of individuals die immediately upon entering the hospital (i.e., if the true delay distribution between hospitalization and death is appreciably bimodal), or if the number of hospital decedents exceeds the number of individuals who are admitted to the ICU. In this sense, our inferred parameter values, e.g., the probability of a patient in critical care dying, should not be interpreted as having real-world meaning, since the values accommodate approximations in order to fit all input data.

Furthermore, by separating the observables from the incidence dynamics, our model supposes that the hospitalization status of an individual does not affect their likelihood of infecting someone. This choice is appropriate if the number of hospitalized individuals is a small fraction of the total number of infected individuals, or if the delay between infection and hospitalization is longer than the serial interval.

The dynamics of the system may be described by daily flux variables:

- *σ_i_*(*t*), the number of infected individuals who become symptomatic
- *h_i_*(*t*), the number of daily admissions to all hospitals
- *r_i_*(*t*), the daily number of patients discharged from all hospitals
- *c_i_*(*t*), the daily number of patients transferred from the main floor of a hospital to its ICU
- *v_i_*(*t*), the daily number of patients transferred from the ICU to the main floor of a hospital, and
- *d_i_*(*t*), the daily number of deaths in ICU rooms.

We do not directly model deaths that happen outside of hospitals but instead infer the ratio of these deaths to the hospital deaths during our fitting procedure, as described below. Figure 1 schematically depicts the topology of our model along with the names of all flux and cumulative variables.

**FIG. 1.**
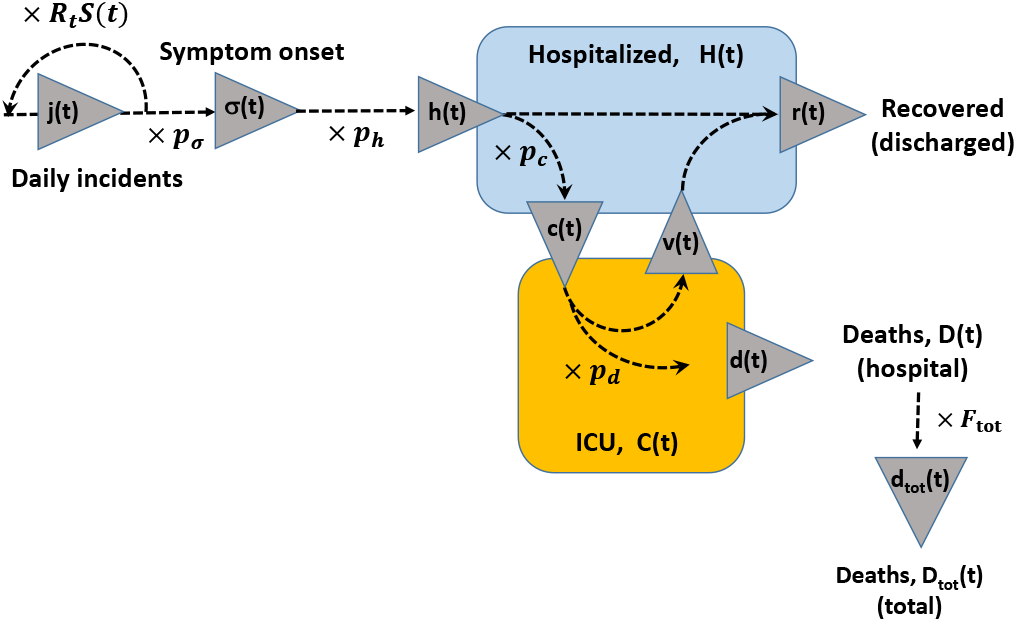
The topology of our model along with the names of all flux and state variables: the daily incidence, *j_i_*(*t*); the daily number of newly symptomatic individuals, *σ_i_*(*t*); the number of daily admissions to all hospitals, *h_i_*(*t*); the daily number of patients discharged from all hospitals, *r_i_*(*t*); the daily number of patients transferred from the main floor of a hospital to its ICU, *c_i_*(*t*); the daily number of patients transferred from the ICU to the main floor of a hospital, *v_i_*(*t*); the daily number of deaths in hospitals, *d_i_*(*t*); and the daily number of deaths in and out of hospitals, *d*_tot,_*_i_*(*t*). State variables are: the total number of occupied hospital beds (main floor) *H_i_*(*t*), and the total number of occupied ICU beds *c_i_*(*t*). The other parameters of the model are the fractions of infected individuals who ever become symptomatic, *p_σ,i_*; the fraction of symptomatic individuals who are ever hospitalized, *p_h,i_.;* the fraction of hospital patients who ever get to ICU, *p_c,i_*; and the fraction of ICU patients who will ultimately die *p_d,i_.;* and the multiplier, *F*_tot_ that converts between hospital deaths and all deaths in the state, including those outside of the hospital system. For the sake of legibility, we suppress age-group indices in the diagram.

The dynamics of any flux variable *y*(*t*) defined above may be obtained from the variable *x*(*t*) directly preceding it in the chain of events shown in Fig. 1:

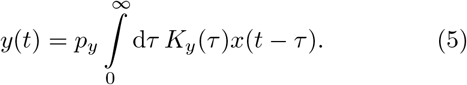

Here, *p_y_* is the proportion of individuals undergoing the transition *x* → *y* with time delays distributed according to a probability density function *K_y_*(*t*). Note that Eq. 2 has the same structure except that both the input *x* and the output *y* variables are given by the daily incidence *j_i_*(*t*).

For the flux variables defined above, one obtains the following equations. The number of infected individuals who become symptomatic is

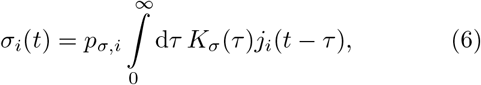

where *p_σ,i_* is the (age-dependent) fraction of infected individuals who ever develop symptoms and *K_σ_* (*τ*) is the PDF of the incubation period. The fraction *p_h,i_* of symptomatic individuals who are ever admitted to the hospital is

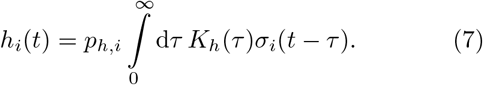

The flux of hospital patients who become critically ill and are admitted to the ICU is given by

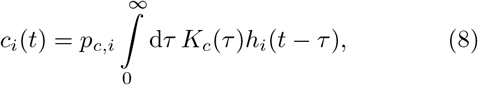

of which a fraction *p_d,i_* ultimately die according to

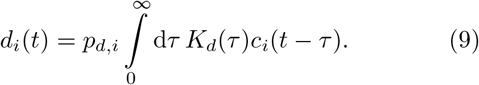

The fraction 1 − *p_d,i_* of ICU patients who stabilize and return to the general ward of the hospital do so according to

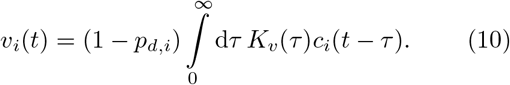

Both stabilized patients and hospital patients who recover without requiring critical care are discharged, thus the influx of recovered individuals due to hospital discharges is given by

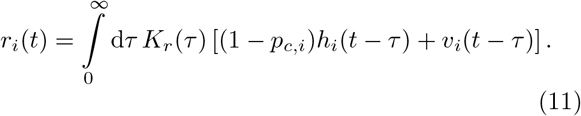

Finally, to approximately account for patients who die outside of the hospital system, we introduce *d*_tot_, a variable that tracks the total number of daily deaths both within and outside of the hospital. We connect total deaths to hospital deaths d according to a prefactor *F*_tot_ ≥ 1 and delay time *τ*_tot_ that reflects bureaucratic delays associated with issuing deaths certificate and publishing data on the Illinois Department of Public Health (IDPH) website. We observe that these bureaucratic delays manifest themselves in strong day-of-the-week effects.

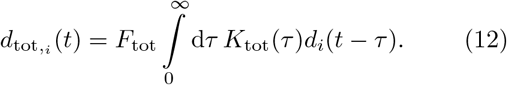

In our simulations, we draw on clinical data to specify the age-dependent rates for hospitalization, ICU admission, and death. We report the details of this severity model in Table S1. To account for differences between the literature values and what has occurred in Illinois, we introduce age-independent prefactors for the transition rates and fit them in our simulations. Although the relative severity values we use may not be accurate, in practice this choice does not affect our model dynamics. Because the contact matrix *ξ_im_* is constant across all interactions, and because the susceptibility is not a function of demographic group, the demographic-aggregated statistics are insensitive to the relative demographic ratios. Then, since we only fit data that has been summed over all age groups, we cannot observe any differences caused by inaccurate severity ratios.

The instantaneous occupation of hospital beds *H*(*t*) and of ICU beds *C*(*t*) may be obtained by integrating the incoming and outgoing fluxes as

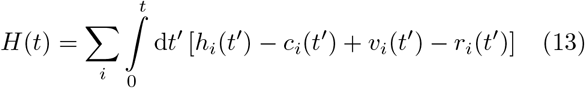

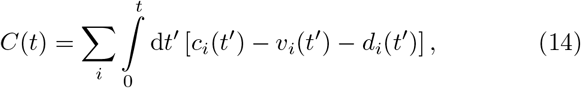

while the cumulative numbers of hospital and total deaths are

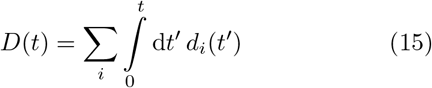

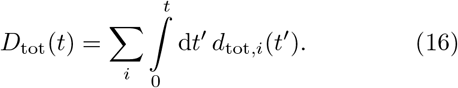

## II. PARAMETER INFERENCE

We calibrate our model to data by sampling over the high-dimensional model parameter space using a Markov chain Monte Carlo (MCMC) approach, as has been done by many others for epidemics in general (see, e.g. [7, 20]) and also for COVID-19 (see, e.g. for applications in China [21], Mexico [22] and Italy [23]). While standard optimization techniques can also identify the best-fit model parameters, we use MCMC because it produces an estimate of the global posterior probability distribution. With the full distribution, we can motivate bounds on parameter uncertainty, explore correlations between parameters, and identify model idiosyncrasies. Access to the full distribution also provides a direct means to marginalize over some modeling uncertainties when forecasting the future trajectory of the epidemic.

### A. Markov chain Monte Carlo methods

Given a set of model parameters 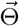 and data 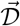, the input to the MCMC sampler comprises a prior 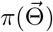 and a likelihood function 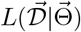. The sampler computes the posterior probability 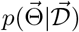 for each point in parameter space according to Bayes’s theorem

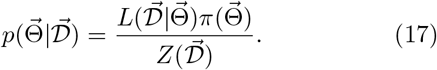

The likelihood *L* represents the probability of observing the data 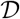 given a model with input parameters 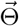, and the prior *π* encodes our expectation for the probability of a given set of model parameters 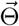. Because we only compare points within the posterior distribution for an individual set of data, we neglect the constant model evidence by setting 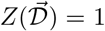. In practice, the MCMC sampling recovers the log of the posterior probability distribution *H* ≡ ln p + const, which combines both type of inputs according to

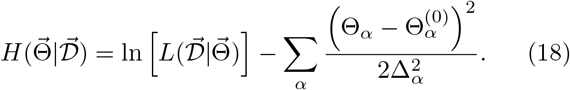

Here, the second term is the log of the prior over the model parameters; for each parameter we either implement a Gaussian prior with mean expected value 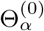 and variance 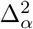, or we use a flat prior in which case the parameter does not appear in the sum.

This Bayesian framework enables a uniform treatment of all available input information information, i.e., both observed time series data and the parameters of the model. We determine the prior means by averaging published clinical data weighted in proportion to the sample size of each study. To account for differences between reported estimates of parameters due to, e.g., possible variability of model parameters between different locations, the tolerance parameters locations, the tolerance parameters were estimated as *unweighted* root-mean-square deviations of the published data from their respective average values 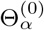 across the different studies. As a result, our procedure is flexible with respect to any local variability in model parameters. If high quality local data on parameter values are available for calibration, as might be the case for the duration of ICU stays or severity models, these data can be used directly with small values (or zero) for the respective tolerance parameters. By forcing parameters with known values to be more tightly constrained, unknown parameters will be automatically optimized with respect to all data types, and we can thus increase the fidelity of our model calibration result.

Many of our model outputs and data quantify daily incidences, e.g., the number of deaths per day. For these sorts of rate data, a Poisson likelihood estimator is appropriate. For a data point *d* at time *t*, the Poisson likelihood is given by

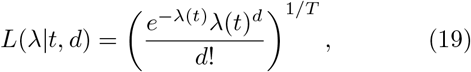

where the time-dependent rate *λ*(*t*) is equal to the model output and *T* is the correlation time for the data. The likelihood over the full data set is the product over the likelihoods for each individual data point; thus, the total log-likelihood is given by

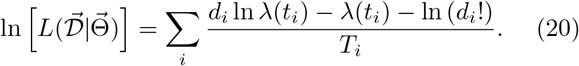

In addition to being a well-motivated choice when comparing count data to rates, the Poisson likelihood is appealing because its effective uncertainty scales with the rate parameter. Thus, unlike with the *L*_2_ norm, a single parameter specifies both the expectation value and the uncertainty of the measurement. In practice, we found that evaluating likelihoods using the *L*_2_ norm did not significantly alter the qualitative features of our forecasts.

We divide by *T* in Eq. 20 because we also calibrate against instantaneous hospital statistics, such as occupancy in the general ward and in the ICU. These data sources have natural correlation timescales: occupancies correspond to smoothed averages since the majority of individuals who occupy a bed do so continuously over several days. We set *T* equal to the correlation timescales Θ^(0)^ from our priors. In particular, we assume a correlation of 6 days for occupied hospital beds, and a correlation of 12.75 days for occupied ICU beds. We set *T* =1 for the raw incidence data, i.e., for daily hospital deaths and for daily total deaths.

In Table I, we enumerate the model parameters we sample over and list the bounds on those parameters’ values. We also describe the shape of any prior distributions we impose. In our model, we use gamma distributions to describe delays, and specify the mean and standard deviation for each distribution. Here, the mean *τ* and standard deviation *σ* of a gamma distribution are related to the standard shape and scale parameters by *k* = *τ*^2^/*σ*^2^ and *θ* = *σ*^2^/*τ*, respectively. We fix the serial interval mean and standard deviation to 4 and 3.25 days respectively [24, 25], while our incubation time distribution has fixed mean 5.5 days and a standard deviation of 2 days [26, 27]. Parameters for all other delays are sampled.

Finally, we implement MCMC sampling using the Python package emcee [33]. To improve sampling efficiency, we use ensemble move proposals based on the differential evolution [34], differential evolution snooker [35], and kernel density [36] proposal updates.

### B. Posterior distributions and data fitting

We calibrate our model using data on hospital and ICU room occupancy by COVID-19 patients, the number of daily deaths of COVID-19 confirmed patients in hospitals, and the total number of daily deaths as publicly reported by the IDPH [37]. At the time of calibration, the hospitalization and ICU data were not publicly available and were provided to us by the IDPH. The MCMC sampling procedure produces a high-dimensional posterior probability. We use this posterior to identify the expectation values and uncertainties for each parameter with respect to the model and the data.

To summarize the posterior distribution in terms of epidemic trajectories, we take a representative sample of parameters 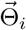 according to their posterior probabilities. For each 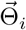, we simulate the full course of the epidemic, and we plot the resulting family of curves in aggregate. At every time point we identify the median values of all measurable quantities (hospital and ICU rooms, and deaths) as well as quantiles corresponding to 68.4% and 95.6% confidence intervals. Because these quantiles are evaluated independently at each point in time, the visually recognizable curves do not correspond to actual epidemic trajectories.

**TABLE I.**
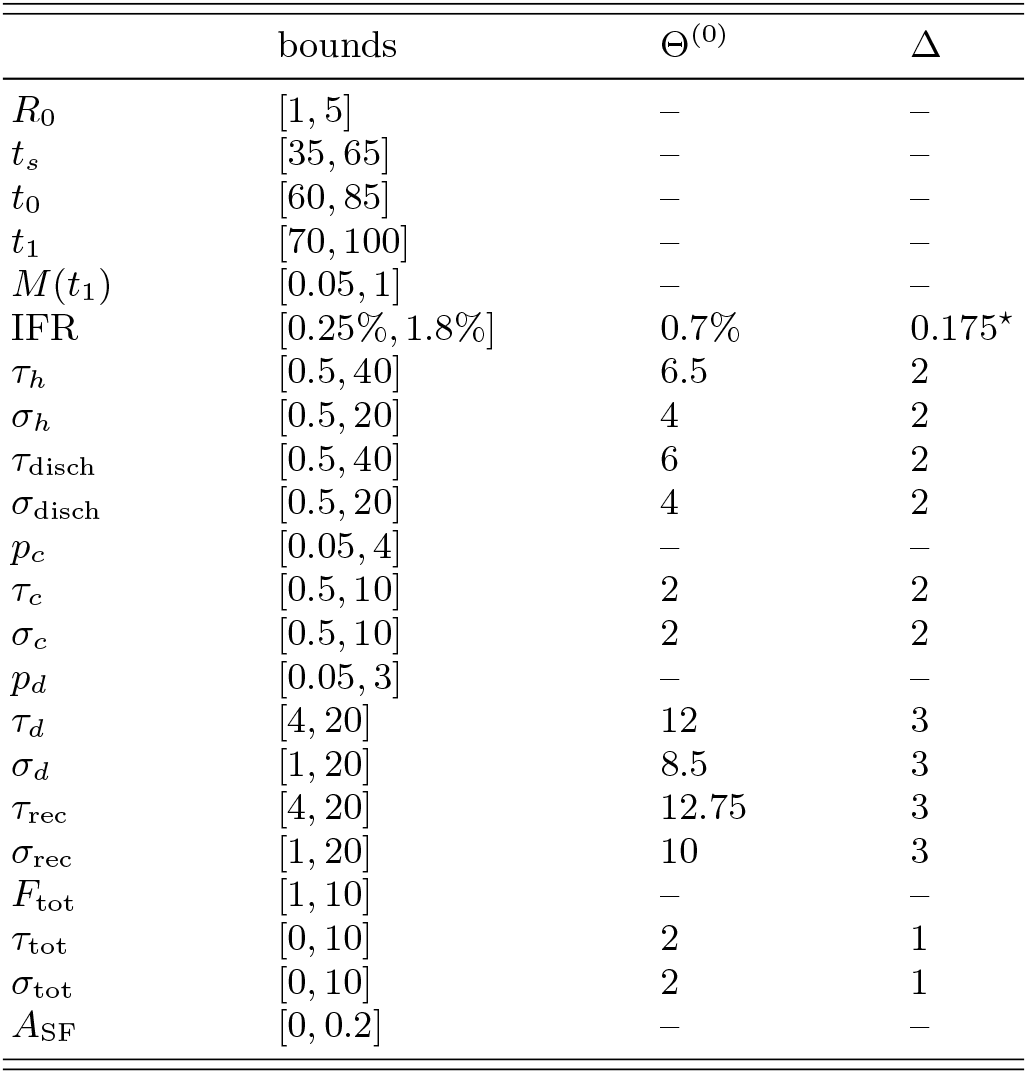
Specification of sampler parameters and their priors. The mean and standard deviation of a delay-time PDF *K_x_* are denoted by *τ_x_* and *σ_x_* respectively, and probabilities *p_x_* denote the overall scaling of the age distribution *p_x,i_* as specified in Table S1. In addition to the listed bounds on the various scaling factors *p_x_*, we also enforce that the number of individuals drawn from a state does not exceed the total number presently in that state. We indicate strict bounds on parameter values and provide the mean Θ^(0)^ and standard deviation Δ for parameters’ Gaussian priors where specified. Our priors on *K_h_*, *K_r_*, *K_c_*, and *K_d_* are informed by Refs. [27-31], and use Ref. [32] to set our prior on ln IFR. *We implement a Gaussian prior for ln IFR, with mean corresponding to IFR = 0.7% and variance of ln IFR set to 0.175.

## III. MODELING RESULTS

We used our age-of-infection model to describe the progression of COVID-19 in Illinois during 2020. We performed analyses for the state and for four distinct localities of the state. We also considered three separate scenarios of social distancing in our modeling.

### A. Simulations for Illinois

In Fig. 2 we show the fits and predictions of our model for the entire state of Illinois assuming that once implemented, the state-imposed and self-regulated social distancing behavior of the population maintains the same level until the end of the simulation. We report the median and the 68.4% and 95.6% confidence intervals of several dynamical outputs of our model, obtained from an ensemble of forward simulations that sample the posterior distribution over model parameters. Fig. 2 presents three separate calibrations: (a) using data up through April 20, 2020 and assuming a fixed seasonal forcing amplitude *A*_SF_ = 0.2, (b) again using data through April 20, 2020 but instead sampling over *A*_SF_, and (c) using data through May 17, 2020 and sampling over *A*_SF_.

**FIG. 2.**
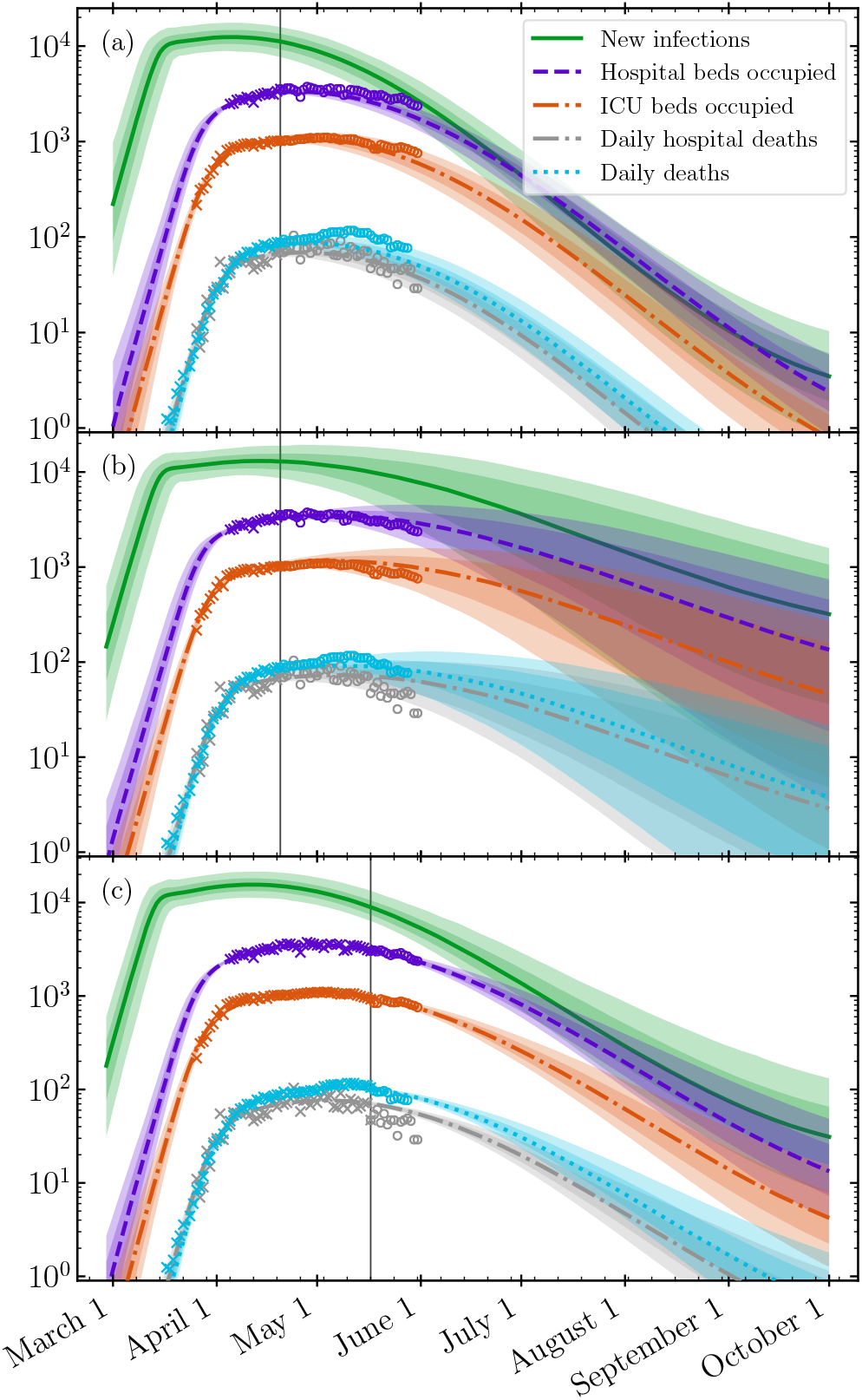
Fits and predictions of our model of the entire state of Illinois under the continuation of the Stay-at-Home order and social distancing measures, resulting from parameter inference which (a) fixes *A*_sp_ = 0.2 and calibrates to data through April 20, 2020, (b) samples *A*_sp_ and calibrates to data through April 20, 2020, and (c) samples *A*_sp_ and calibrates to data through May 17, 2020. In each case, the data used for calibration is denoted by crosses, while data from later dates (open circles) allow a comparison of model predictions with subsequent real world observations. Each panel’s vertical line marks the end of data used for calibration. Solid curves denote median model predictions at a given time, while the shaded regions denote the 68.4% (darker shading) and 95.6% (lighter shading) confidence intervals of a particular output. We depict the daily incidence of new infections (green, solid), the number of occupied hospital beds by confirmed or tentative COVID-19 patients (purple, dashed), the number of ICU beds occupied by confirmed COVID-19 patients (orange, dot-dashed), the number of hospital deaths by confirmed COVID-19 patients (grey, dot-dashed), and total daily deaths of COVID-19 patients (light blue, dotted). We remove reporting artifacts by plotting daily death data smoothed by a 7-day running average.

First, comparing panels (a) and (b) demonstrates that a 20% seasonal forcing effect produces a worse projection and may be an overestimate; while our model does not infer the origin of any yearly, periodic modulation to *R_t_* (or lack thereof), by mid May the forecasts disagree with the data. By contrast, the uncertainty introduced by sampling *A*_SF_ produces a more robust fit to the long plateau exhibited in all data sources. Next, comparing panels (b) and (c) shows that while short-term predictions are broadly consistent, the spread of model forecasts become narrower. In particular, including data up to May 17, as in panel (c), enables the model to identify that the plateau is beginning to bend, in contrast to the model shown in panel (b), in which a continued plateau is not precluded. We investigate the predictive power of our model and calibration procedure in the next subsection.

Hospital and ICU occupancy as well as deaths related to COVID-19 exhibit a long plateau spanning from the beginning of April through at least mid May. This behavior is not just due to the fact that mitigation reduced *R_t_* to almost exactly one, but also because of the variance in when the lagging indicators respond to the rate of infection. Namely, while the various non-pharmaceutical interventions (NPIs) implemented in Illinois occurred on relatively short timescales (as can be seen in the sudden change of slope for the daily incidence of new infections in Fig. 2), the variance in the delays between when individuals become, e.g., symptomatic and then hospitalized, introduces spread in the indicators’ responses to changes in *R_t_*. As an example, some portion of individuals infected before any mitigation takes place will continue to be admitted to the hospital well after mitigation occurs. Indeed, this variability compounds with subsequent transitions, such that daily deaths, being the final indicator, exhibit the most gradual change in incidence rate. Our model’s generality to arbitrary delay distributions makes it particularly well-suited to accurately capture this effect.

### B. Robustness of predictions

In order to explore the predictive capabilities of the model, we present a series of benchmark simulations to compare the predictions of models calibrated with increasingly recent data. In Fig. 3, we show the fits to hospital beds occupied, ICU beds occupied, daily hospital deaths, and daily total deaths in the entire state of Illinois, calibrating with data up to April 1, 2020, April 8, 2020, April 20, 2020, and May 3, 2020. During the time period studied here, the Illinois Stay-at-Home order was issued, leading to an end to the exponentially growing phase of the epidemic and a flattening of the curve. Due to transition delays, the quantities to which we calibrate exhibit exponential growth as late as early April; for example, the number of daily deaths does not flatten until around April 10, 2020. Thus, the above-specified dates of calibration provide a test that measures the ability of our model to anticipate the bending of the curve.

**FIG. 3.**
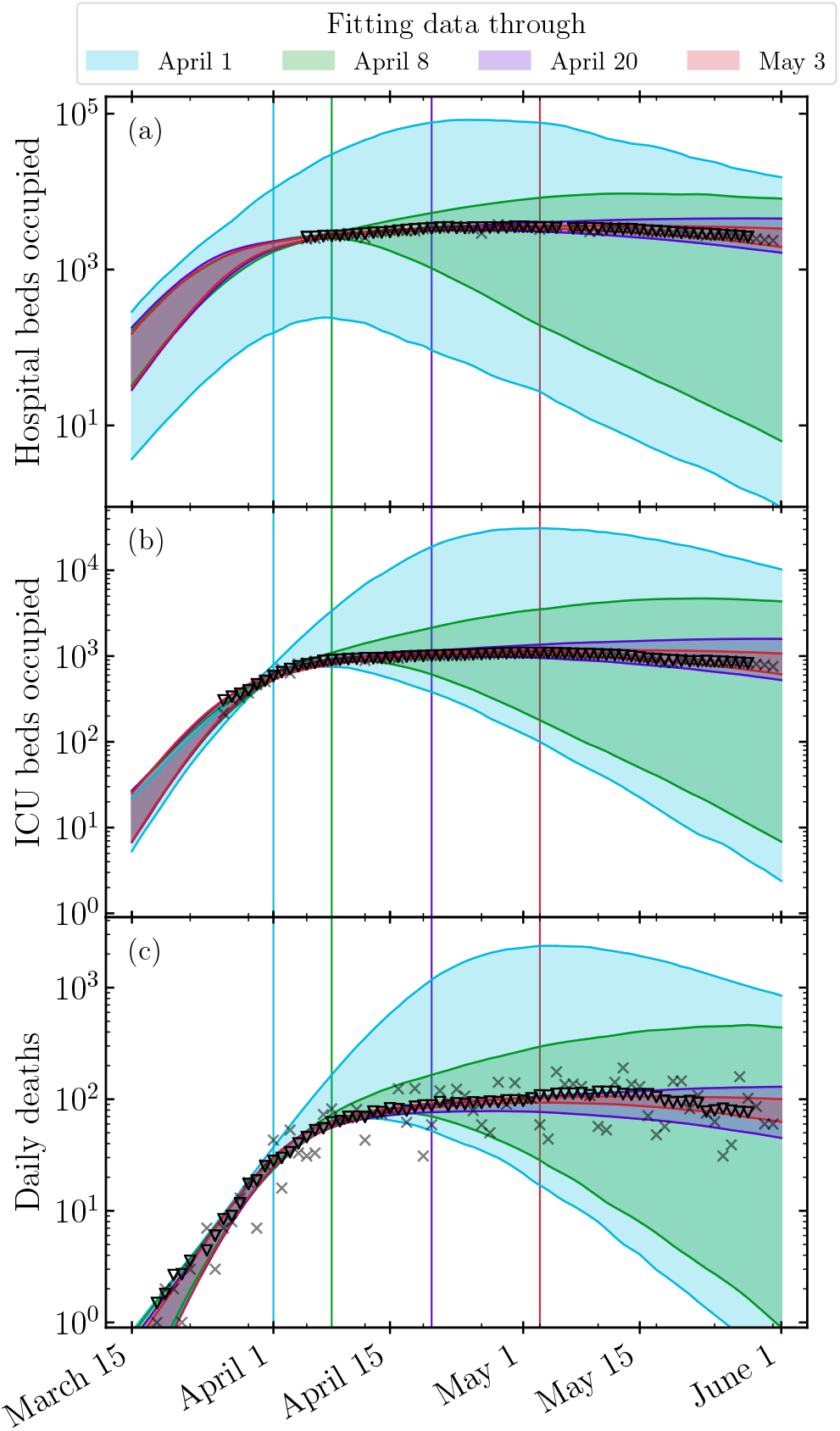
Robustness of simulations for the COVID-19 epidemic in Illinois, evaluated by comparing 95.6% confidence regions of predictions for parameter inference calibrated to data through April 1 (light blue), April 8 (green), April 20 (purple), and May 3, 2020 (red). The panels depict the number of hospital beds occupied (a), the number of ICU beds occupied (b), and the total daily deaths (c). In each panel, triangles depict the data smoothed by a 7-day rolling average and grey crosses the actual data.

As expected, simulations with earlier terminal calibration dates generate forecasts with larger, less restrictive confidence intervals: with data from only the earliest stages of the epidemic, neither continued exponential growth nor successful suppression of infections due to mitigation can be ruled out. Between April 1 and April 8, however, ICU occupation stopped growing exponentially, thereby providing the first indicator by which the model can infer the effect of mitigation. Indeed, even the April 8 forecast anticipates the subsequent flattening in the daily deaths curve. Later forecasts (e.g., that of April 20) remain consistent with the April 8 model, while also favoring a continuation of the plateau over a more rapid decline in use of hospital resources and deaths. Finally, the May 3 model is largely consistent with the April 20 one, but begins to project a slight decline that agrees with the new data. This latest forecast should not be expected to lie strictly within the confidence interval of the previous, as any future changes in mitigation cannot be anticipated.

We also point out that the April 1 forecast for hospital occupation spans several orders of magnitude on any given day (as seen in panel (a) of Fig. 3). This is due in part to the lack of hospitalization data that could provide direct constraints, but is also an inevitable feature of a system which exhibits exponential growth dynamics. Furthermore, we hypothesize that the lack of hospitalization data before the beginning of the plateau is at least partially responsible for the April 1 forecast for ICU occupation and daily deaths being relatively unconstrained. Since hospitalizations serves as the earliest available indicator for the progression of the epidemic, we hypothesize that if it were supplied with this data before April 1, the model would have been able to discern that the data were no longer consistent with an exponentially-growing epidemic. This observation underscores the importance of rapid and reliable reporting of hospitalization data—or even better, robust and representative testing for positive cases— in the context of modeling epidemic dynamics.

The disappointingly short horizon of predictability for epidemic models when *R_t_* > 1 shown here represents a fundamental limitation of forecasting, in much the same way that chaotic dynamics limits weather prediction, and this issue has been noticed in other independent studies [38-40]. However, the exponential sensitivity has a silver lining: small changes to transmission can have large impacts on the overall trajectory and fatality of the disease.

In summary, the model curves fitted after April 8, 2020, i.e., with data from the plateau, follow the trends of the data well. We conclude that the model can be characterized as semi-quantitative and that it is capable of capturing broad epidemic dynamics and fitting empirical data. In this sense, it can serve as a useful tool to make short term predictions that may be useful for planning purposes.

### C. Regional modeling

To account for differences in the epidemic in different parts in the state, we simulate the epidemic trajectory in the four distinct Restore Illinois regions separately. Each of these “super-regions” is composed of multiple of the Emergency Medical Services (EMS) regions defined by the Illinois Department of Public Health (IDPH) [41]. The Northeast super-region includes the city of Chicago and its suburbs, EMS regions 7 through 11. The North-Central super-region contains EMS regions 1 and 2, the Central super-region EMS regions 3 and 6, and finally the Southern super-region comprises EMS regions 4 and 5. In the absence of detailed transportation data, we assume no population transfer between these super-regions, and so unlike, e.g., the model for the state of Georgia in Ref. [42] and Italy [23], our regional modeling is not a genuine metapopulation model for the state.

In Fig. 4 we show the fits and predictions of our model calibrated to data up to May 17, 2020 for each of these four regions. We report inferred model parameters in Table S2. Our median estimates of the basic reproduction number *R*_0_ at the start of the epidemic are consistently above 1, ranging from 2.4 ± 0.16 for the Northeastern region including Chicago and its suburbs to 1.7 ± 0.16 for the Central region including the University of Illinois at Urbana-Champaign.

**FIG. 4.**
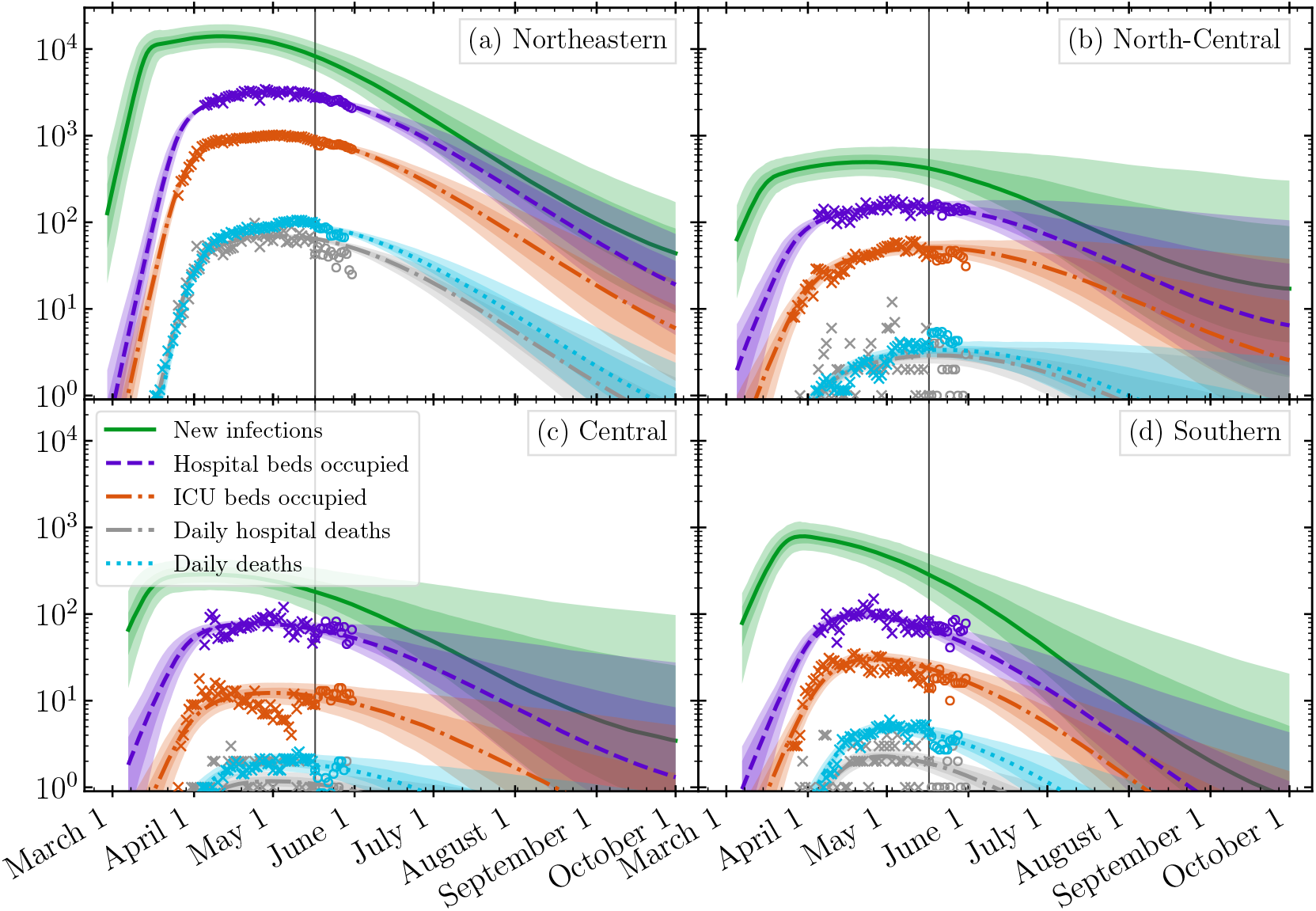
Fits and predictions of our model under the baseline scenario with the Stay-at-Home order and social distancing maintained until October 1, 2020. The model is calibrated with data (crosses) through April 20, 2020 separately for each of the four super-regions of the state: (a) North-Central, (b) Northeast, (c) Central and (d) Southern. The data from later dates (open circles) were not used in our fits and allow a comparison of model predictions with real world observations. Solid lines of different denote the median model prediction at a given time. The shaded regions denote the 68.4% (darker shading) and 95.6% (lighter shading) confidence intervals, obtained as quantiles of an ensemble of forward simulations which sample the posterior distribution over model parameters. We depict the daily incidence of new infections (green, solid), the number of occupied hospital beds by confirmed or tentative COVID-19 patients (purple, dashed), the number of ICU beds occupied by confirmed COVID-19 patients (orange, dot-dashed), and total daily deaths of COVID-19 patients (light blue, dotted). We remove reporting artifacts by plotting daily death data smoothed by a 7-day running average.

The per capita daily deaths and illnesses is at least ten-fold higher in Chicago and its suburbs compared to the downstate areas of Illinois. This is likely due to increased contact density in the upstate region, reflected by a higher initial *R*_0_ ≈ 2.3 compared to ≈ 1.8 in the downstate regions, coupled with the fact that mitigation began earlier relative to the start of the epidemic in some downstate regions. In regions where the virus entered the population later, the epidemic had a shorter phase of unmitigated exponential growth.

The values of *R_t_* corresponding to the post-mitigation basic reproduction number are very close to 1, reflecting the approximately constant number of hospital and ICU beds occupied by COVID-19 patients and daily deaths from COVID-19 in different super-regions in Illinois during much of April and May 2020.

### D. Evaluation of parameter fits

In Fig. 5, we show a subset of the joint posterior probability distributions for model parameters relevant to the parameterization of the mitigation factor *M*(*t*) as specified above, fitting to data shown in Fig. 2. The correlations between some pairs of fitted parameters, e.g., between *R*_0_ and the start date of the epidemic t_s_, are reflected in the ellipsoidal shape of the posteriors. This is sensible: the later the epidemic begins, the larger the basic reproduction number must be in order to fit the data.

**FIG. 5.**
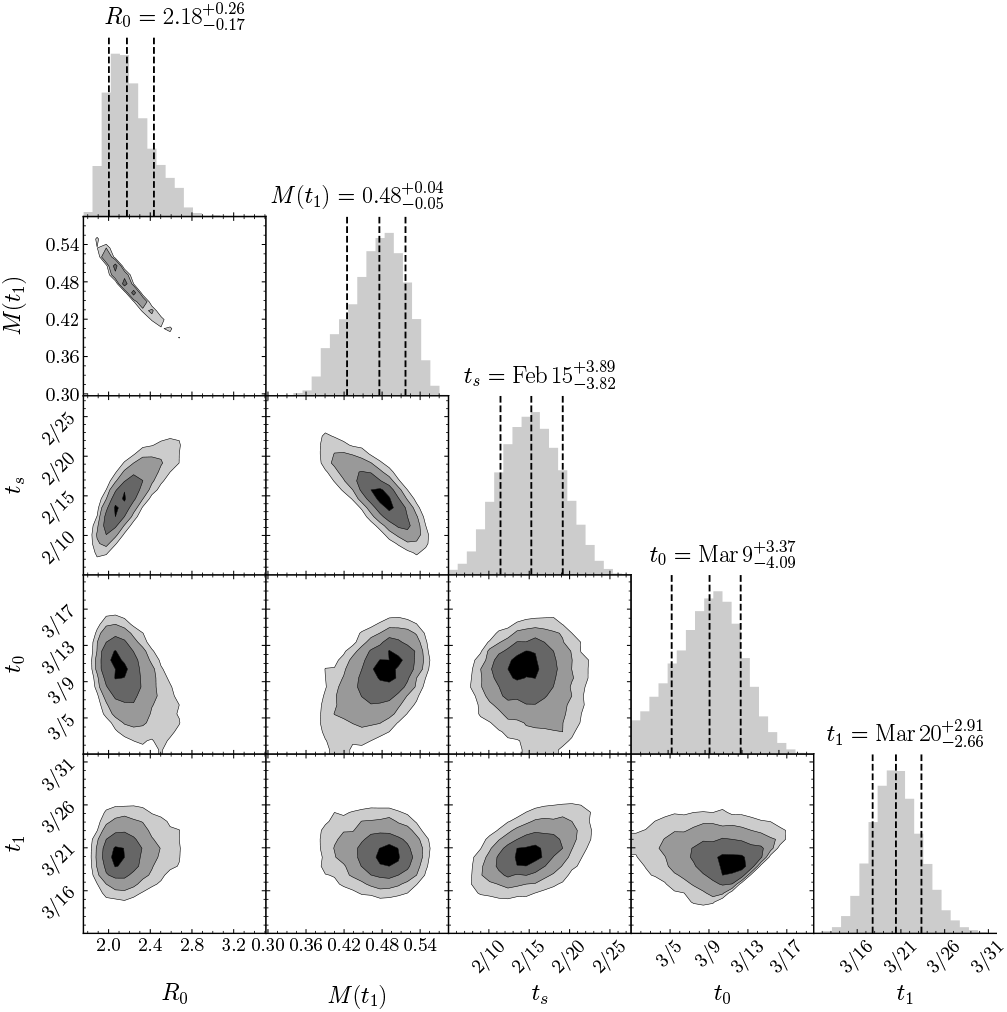
Joint posterior distributions of pairs of the main parameters of our model fitted to the all-state data in Illinois up to May 17, 2020. The variables shown are the initial value of the reproduction number, Ro, the mitigation factor *M*(*t*_1_), the start date of the epidemic *t_s_*, the day the mitigation factor begins to deviate from 1, *t*_0_, and the day the mitigation reaches its asymptotic value, *t*_1_.

In Table S2, we report the parameter values our model infers when fitting to data for different regions of the state and over different time ranges. We also report the effective reproduction numbers *R_t_* as evaluated on May 1, 2020. On May 1, 2020, *R_t_* appears to have barely dropped below unity, suggesting that mitigation efforts may have only marginally halted the exponential growth of the epidemic at that time.

### E. Comparison with mobility data

While our model does not provide a microscopic description of social interactions and movement in the population, we may evaluate our fitted *M*(*t*) relative to social mobility indices derived from cell phone data [43, 44]. In the top panel of Fig. 6, we plot the time dependence of several mobility indices reported by Google [43] for the entire state of Illinois, measuring change in visits to destination points categorized as retail and recreation, grocery and pharmacy, parks, transit stations, and workplaces. The Unacast mobility data [44] is shown in bottom panel of Fig. 6, depicting an effective distancing metric and a measure of trips to so-called non-essential destinations.

**FIG. 6.**
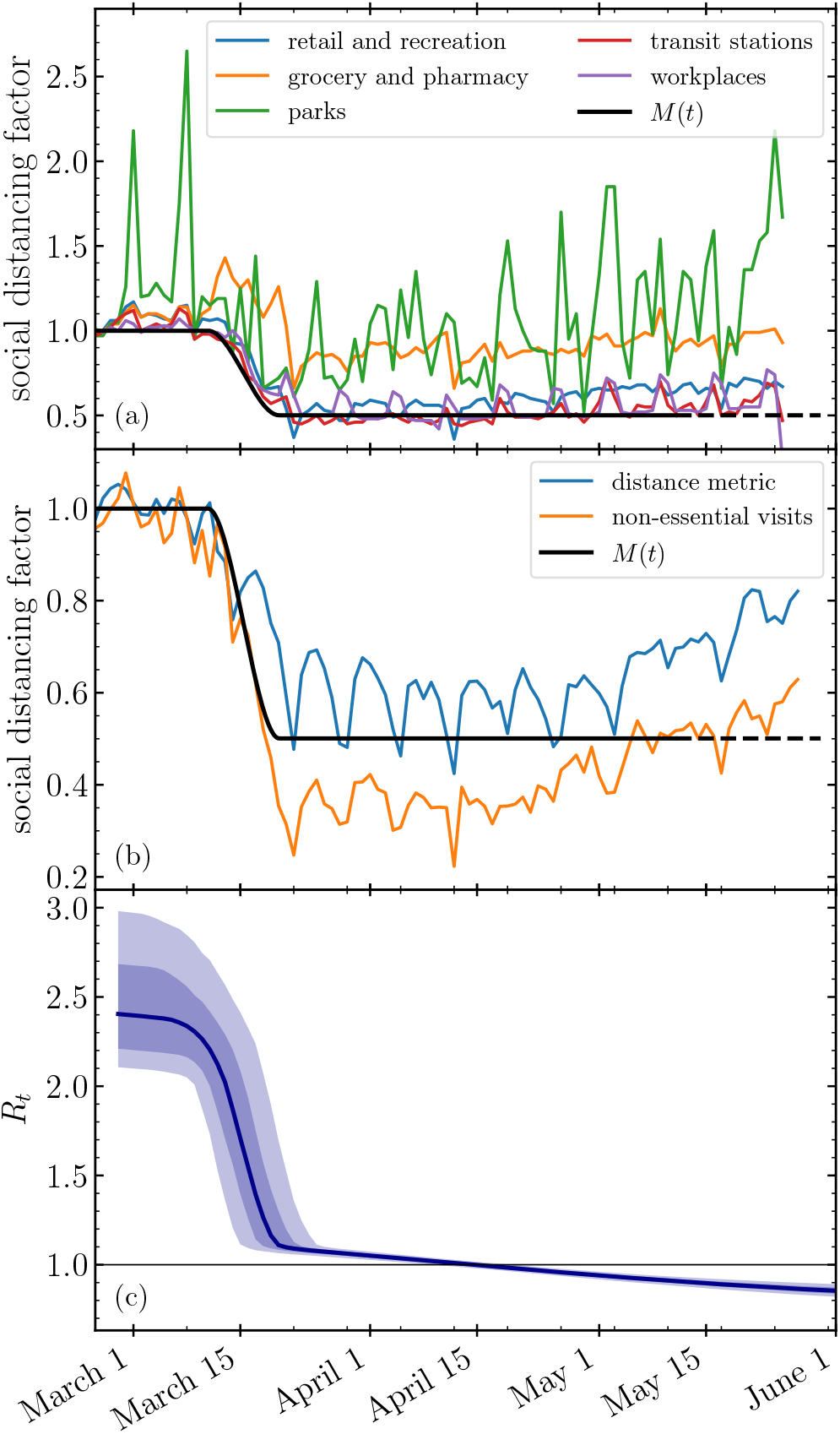
The time-dependent mitigation factor *M*(*t*) defined in Section IA as inferred from model fits compared to measures of mobility in Illinois provided by Google (a) and Unacast (b). In (c), we also plot the inferred evolution of *R_t_* as defined in Eq. 3, which we draw from a calibration to data from the entire state of Illinois through May 17, 2020. Shaded regions denote the 68.4% (lighter shading) and 95.6% (darker shading) confidence intervals, while the solid curve denotes the median.

Remarkably, although the model is supplied with no prior information on non-pharmaceutical interventions, the inferred dates and magnitude of mitigation agree with the start and end dates of the largest drop in both sets of mobility data. Comparing to the Google mobility data, our *M*(*t*) curve exhibits an amplitude, start date and end date consistent with indices corresponding to retail and recreation, transit stations and workplaces. On the other hand, parks and grocery and pharmacy show a more modest reduction which still matches the time frame of *M*(*t*). In the Unacast data, both metrics also appear to match the time-dependent change in *M*(*t*). Note that both datasets evince increased movement at later dates. Because we here parameterize mitigation as a single transition, our fits to *M*(*t*) would not reproduce this recent increasing trend.

Our procedure is distinct compared to several previous efforts to incorporate mobility into models of COVID-19 dynamics, which either impose that changes in transmission coincide with known dates of non-pharmaceutical interventions, or scale *R_t_* according to reductions in mobility [45, 46]. The fact that our model independently recovers a trend in mitigation consistent with mobility measures speaks to its flexibility and calibration procedure.

## IV. WHAT-IF SCENARIOS

We now consider possible future scenarios and alternative historical scenarios in which non-pharmaceutical interventions are lifted or were never implemented at all. The former enables a model-based assessment of the risk of, e.g., lifting Stay-at-Home orders on a certain date, while the latter demonstrates the impact that previously-implemented measures have already had on outcomes. Our analysis focuses on two key measures for guiding and justifying policy decisions: the stress imposed on the healthcare system and the death toll.

### A. No Stay-at-Home order

We first investigate the trajectory of the COVID-19 epidemic in Illinois in the absence of any form of social distancing or mitigation measures, whether self-imposed or mandated by the government. We conclude that rate of hospital-bound deaths, ICU bed occupancy, and hospital bed occupancy would be higher than what actually took place by an order of magnitude or more, as shown in Fig. 7. The Stay-at-Home order and self-imposed social distancing measures were clearly crucial to flattening the curve.

**FIG. 7.**
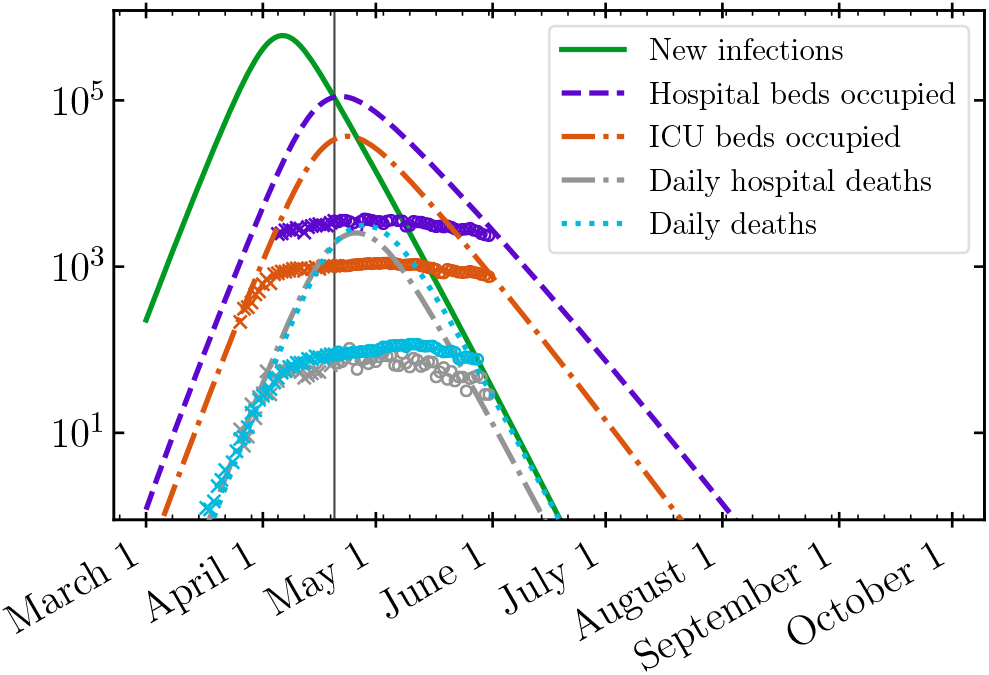
Counterfactual simulation of the COVID-19 epidemic in the absence of government or population self-imposed social distancing measures. The model is calibrated to data through April 20, 2020 (crosses); we then artificially set *M*(*t*_1_) = 1, i.e., assume no mitigation ever takes place. The data from later dates (open circles) are not used in our fits and demonstrate the effect that real-world NPI strategies had on the epidemic. We depict the daily incidence of new infections (green, solid), the number of occupied hospital beds by confirmed or tentative COVID-19 patients (purple, dashed), the number of ICU beds occupied by confirmed COVID-19 patients (orange, dot-dashed), and total daily deaths of COVID-19 patients (light blue, dotted). We remove reporting artifacts by plotting daily death data smoothed by a 7-day running average.

In Ref. [47] we made an early estimate of the ICU utilization in the city of Chicago under two scenarios: one in which the Stay-at-Home order was issued on March 20, 2020, and another in which the order was delayed by 20 days. Under the first scenario, the ICU utilization by COVID-19 patients never exceeded the number of ICU beds not occupied by other patients, while under the second scenario it exceeded ICU capacity by nearly a factor of ten. This example highlights the cost of mistiming in NPIs [48]. In spite of inevitable uncertainties associated with these early estimates, that work correctly identified the timing of the peak in ICU demand to happen on or around April 22, 2020. However, the magnitude of this peak was underestimated in this study, because the scenario assumed that post-mitigation value of *R_t_* = 0.9 would be achieved by social distancing. In reality, the response of the population to the Stay-at-Home order in Chicago was somewhat weaker resulting in a larger value of *R_t_* and about a three-fold higher peak ICU bed occupancy than we had predicted.

### B. Partial removal of Stay-at-Home order

The state of Illinois lifted its original Stay-at-Home order on May 30, 2020 [1]. In Fig. 8 we consider two scenarios for the lifting of Stay-at-Home orders for the entire state of Illinois. In the first scenario, mitigation effects are completely removed and *M*(*t*) = 1 for times *t* after June 1, 2020. We also consider the more conservative case that mitigation recovers by 30% to *M*(*t*) = *M*(*t*_1_) + 0.3(1 − *M*(*t*_1_)) for *t* after June 1, 2020. This second scenario assumes that a combination of self-regulation and remaining government-imposed mitigation measures, such requiring wearing masks, banning large gatherings, etc., results in only a partial reduction of the effective mitigation factor.

**FIG. 8.**
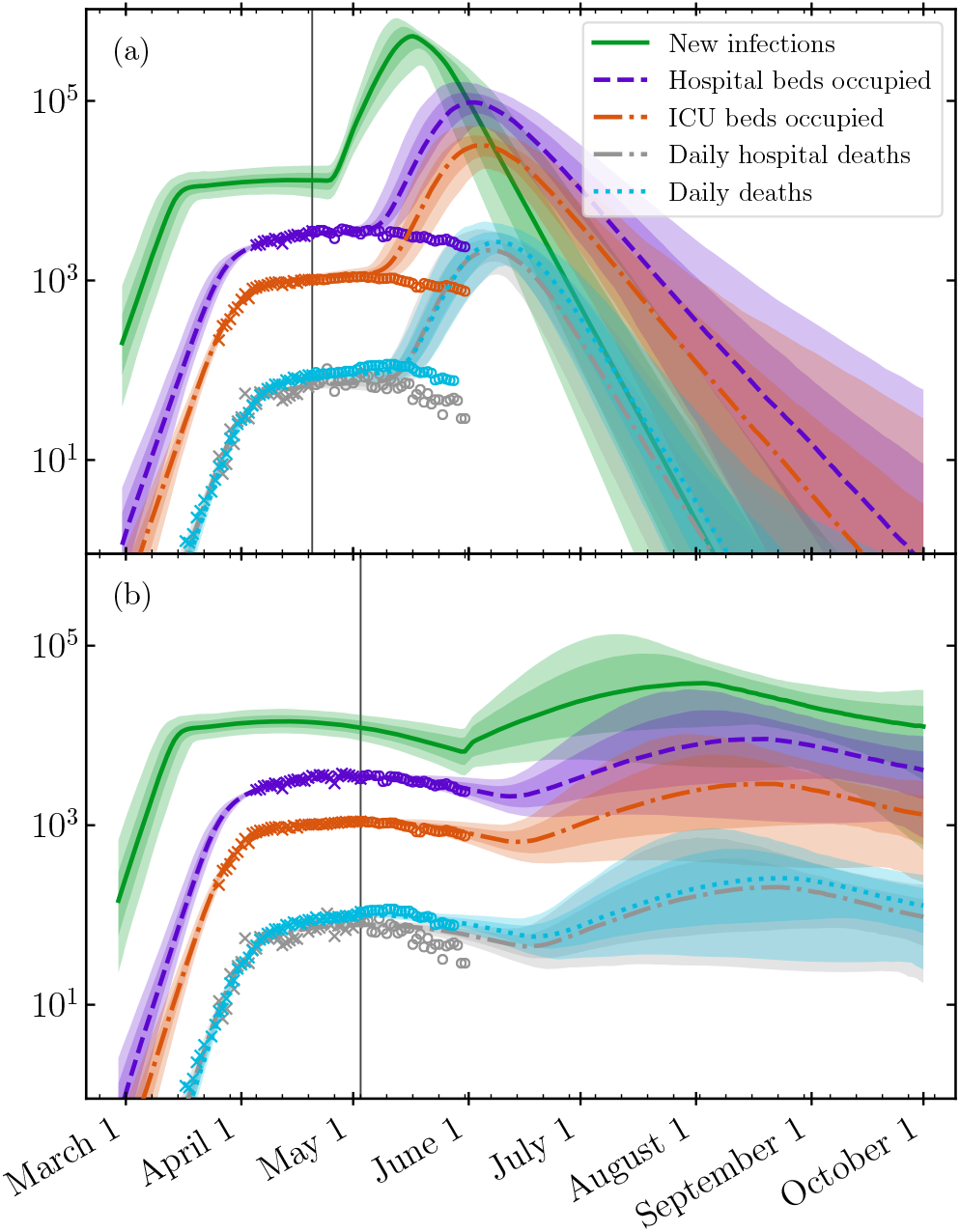
Simulation of different scenarios for relaxation of the Stay-at-Home order for the entire state of Illinois. Each panel’s vertical line marks the end of data used for calibration. We display two scenarios: (a) where the mitigation actor *M*(*t*) returns to 1 on April 24, 2020, corresponding to a complete lack of social distancing, and (b) where on June 1, 2020 the effect of NPIs is reduced by 30%.

The first scenario exhibits a substantial second wave, with rapid and strong peaks in all quantities occurring successively through the month of July. In the second, weaker (and perhaps more realistic) scenario, a second wave still occurs but does so later and with a reduced peak height. In this case, the peak demand for hospital and ICU beds and number of deaths are reduced.

An aggregated model of the entire state does not describe its heterogeneous population structure, which may be particularly important in forecasting beyond the lifting of the Stay-at-Home order. We now report separately the results of modeling the expiration of the Stay-at-Home (with a 30% reduction in mitigation) in each of the four aforementioned super-regions, with the caveat that we are unable to take into account possible transfers of people between the regions. As in Fig. 9, we first calibrate models using data through May 17, 2020, imposing again that mitigation recovers by 30% to *M*(*t*) = *M*(*t*_1_)+0.3(1 − *M*(*t*_1_)) for *t* after June 1, 2020.

**FIG. 9.**
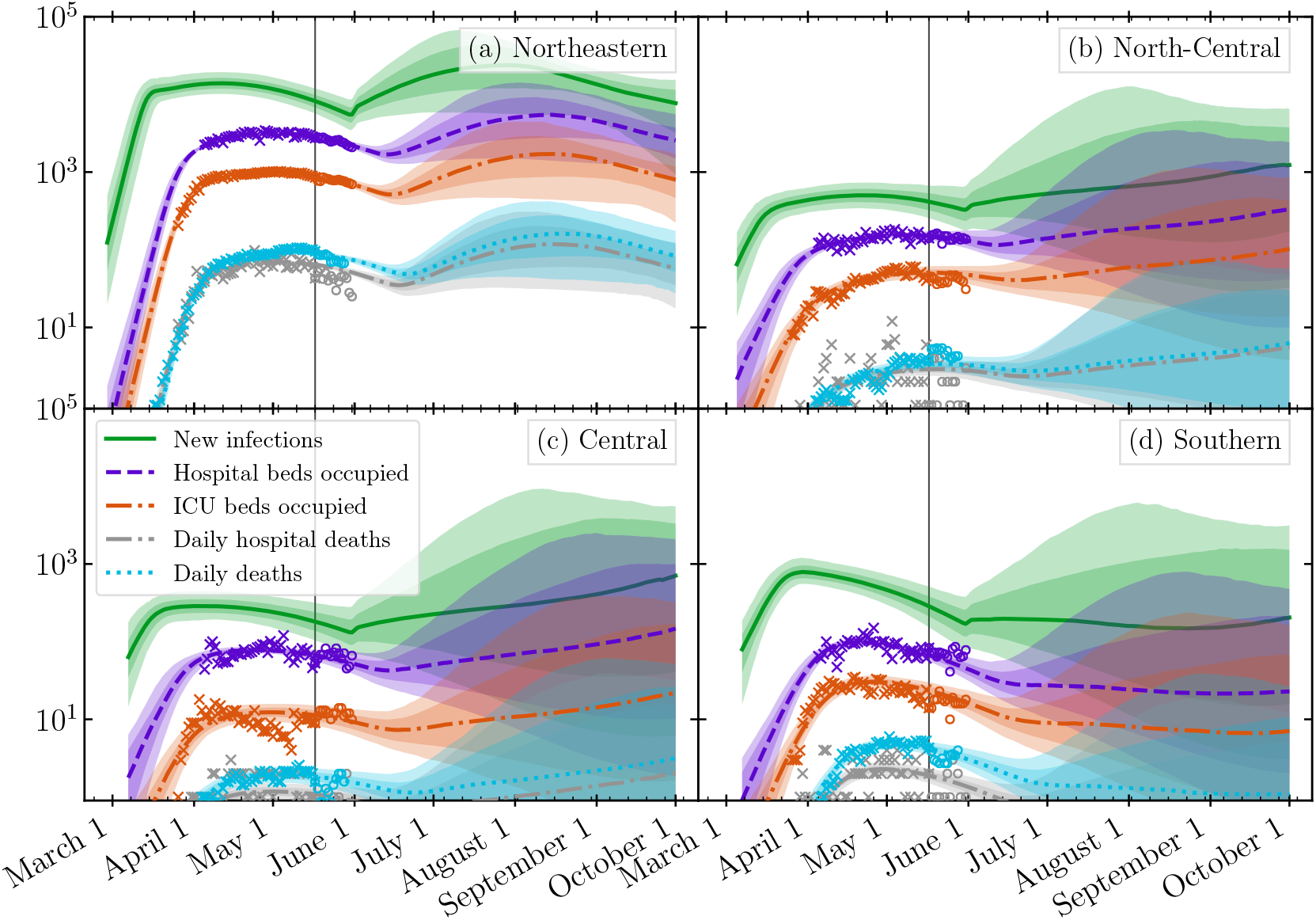
Fits and predictions of our model under the a partial release of the Stay-at-Home order on June 1, 2020 to a mitigation factor *M*(*t*) = *M*(*t*_1_) + 0.3 *×* (1 − *M*(*t*_1_)). The panels and and plots denote the same regions and data sources as those in Fig. 4.

In each super-region, a second wave is clearly visible, as all four populations still today have a significant number of infectious people at large. Although the Northeast region, which includes Cook County and metropolitan Chicago, has the biggest second wave in absolute numbers, its relative impact on the epidemic trajectory is smaller merely because the epidemic has not declined to the same extent as the other three regions. Under the worst case scenario, the more sparsely populated Southern, Central, and North-Central regions have relatively larger second waves because the epidemic as of yet is relatively less active in those regions compared to the Northeastern region, which includes Chicago and its suburbs.

We note that the magnitude of the second wave may be containable if rapid and efficient testing, contact tracing, and isolation mitigation strategies are employed [49, 50]. Following standard protocols [51], we estimate that even if purely manual contact tracing is employed for all three stages of tracing (case identification, tracing, and follow-up), the number of contact tracers required is approximately 8.3 times the daily number of new cases identified. A considerable reduction in workforce and an increase in efficiency can be obtained by electronic measures. Nevertheless, the potential magnitude of the second wave in this scenario suggests that contact tracing will be challenging and require extraordinary resources to execute.

## V. DISCUSSION

Modeling plays an important role in the societal response to the COVID-19 pandemic, and a variety of approaches are used to inform public health policy, guide resource allocation, and plan non-pharmaceutical interventions [52]. The present study of the spread of COVID-19 in Illinois reveals both the strengths and limitations of modeling, and provides potentially actionable insights into the future spread of the disease. We begin with some technical points and best practices that we have developed during our work.

### Model calibration

Our analysis highlights the importance of choosing appropriate data with which to calibrate models, and to perform calibration with precision. Due to the large number of parameters that inevitably enter epidemiological models, calibration requires parameter inference in a high dimensional space with strong potential for improper fits resulting from failure to reach global optima. Although the MCMC methods we use are computationally intensive, they are relatively efficient in exploring high(a) Northeastern dimensional, multi-modal distributions, and converge to well-behaved global posterior probability distributions. Bayesian inference enables the incorporation of previous studies (e.g., meta-analyses) to provide reasonable priors on parameters which are poorly constrained by the available data. As an example, although the data we calibrate to does not constrain the prevalence of the infection, we systematically account for this uncertainty by informing our prior on the infection-fatality rate (IFR) from serological studies [32]. The IFR is an important variable in terms of disease outcomes, and so model predictions must systematically account for the uncertainty in this variable.

In our analysis, we have taken data at its face value. A more thorough analysis could account for differences in the trustworthiness of data, e.g., programmatically dealing with uncertainties associated with classification of early hospital admissions as either COVID-positive or under investigation. Such an approach might also be able to consider the number of individuals who test positive for the virus. Furthermore, because we do not explicitly treat underreporting, data from early in the epidemic may be biased low.

### Error estimation

The MCMC procedure generates a complex, high dimensional probability distribution that may be sampled to estimate future epidemic trajectories. While one might be tempted to simply generate the maximum likelihood trajectory for forward (in time) simulation, this can be misleading. The probability distribution of trajectories may have a maximum likelihood trajectory that is favored only slightly above other trajectories yet is noticeably different from the mean or median estimator. We thus sample trajectories forward in time, and at each time point we evaluate the median and confidence intervals of outputs. Although the curves we depict do not correspond to a trajectory that would be realised in practice, together with confidence intervals, they provide a meaningful description of the range of forecast results.

### Robustness

In order to ascertain the predictive power of our model, we assessed to what extent it was able to make predictions beyond the range of data to which it had been calibrated. We first considered a rather severe test: how far in advance can the model predict the abrupt flattening of the epidemic curves that occurred after April 10, 2020? Unsurprisingly, the answer here was not particularly encouraging because the curves dramatically switch from exponential growth to a phase of much slower variation or plateau. We were only able to fit daily death and ICU occupancy data before the flattening, since hospitalization data was not available until mid-April. As a result, none of the information feeding into the model dynamics were able foreshadow the onset of the plateau.

Nevertheless, such self-consistency checks are important validations of the modeling process, and should be attempted if there are early enough data available. Encouragingly, the range of our model estimates decreased in a hierarchical and consistent manner, with projections from earlier points in time bounding later projections. Of course, it is not possible for our model to predict future changes to the strength of mitigation. Furthermore, our mitigation model only implements a single event and thus cannot account for future short term changes. Real world considerations, such as holidays and quarantine fatigue, would be inconsistent with our mitigation model approximation on long timescales. As such, we do not necessarily expect future predictions (which could account for future changes in mitigation) to lie within the confidence bounds of older ones.

### Correlation with mobility data

For policy and planning purposes, it is important to evaluate the effectiveness of various measures in mitigating the epidemic relative to their, e.g., economic costs. Although epidemiological modeling can in principle provide insight into the former issue, doing so would require a more fundamental description than the one presented here, for example making use of agent-based models [12, 13, 16] or employing spatially-structured and heterogeneous network descriptions [15]. Mean field models, which assume a well-mixed population, do not capture the microscopic effects that could be correlated to specific mitigation strategies.

Nevertheless, mobility data present a potential means to guide and evaluate which policies and social interventions lead to the strongest reduction in disease transmission. While our treatment does not provide such a description or analysis, the correlation between the effective measure of the impact of NPIs, *M*(*t*) (see Eq. 3), and mobility data presented in Fig. 6 is striking. Note that in contrast to Ref. [46], we do not use the mobility data as an input to our calculations. Although our model makes no claims about the effect of changes in the reported mobility indices, this observed correlation encourages more detailed modeling to evaluate the impact of different social distancing measures. Furthermore, our results support the utility of anonymized, aggregated mobility data as a potential low-latency measure of the impact of, e.g., government-mandated social distancing measures on actual population movement, especially since such indicators may provide a predictor of the measures’ influence on epidemic dynamics. A robust understanding of the mechanistic relationship between mobility measures and the spread of the infection could also guide efficient testing and contact tracing strategies, for example using risk-based surveillance methods [53, 54].

### Model generality

Although we designed and tested our model using data for the state of Illinois, the model is general and can be fit to other geographical regions as well, as long as there is availability of hospital, ICU and death data. Using the procedure described above, we calibrate model parameters to public data for hospital and ICU bed occupancy and daily deaths in the New York City data published by the New York City Department of Health and Mental Hygiene [55-57]. We present the results of the fit in Figure 10. With no modification, the model generates a forecast that is remarkably consistent with subsequent data. These fits are subject to the stated limitations of our modeling. In particular, future changes in voluntary and state-mandated mitigation are of course not included. Furthermore, there are possible indications of strong population heterogeneity and overdispersion in the unexpected smallness of *τ*_disch_ compared to the value for Illinois. A detailed discussion of these effects is beyond the scope of this paper. We report model parameter values in Table S3.

**FIG. 10.**
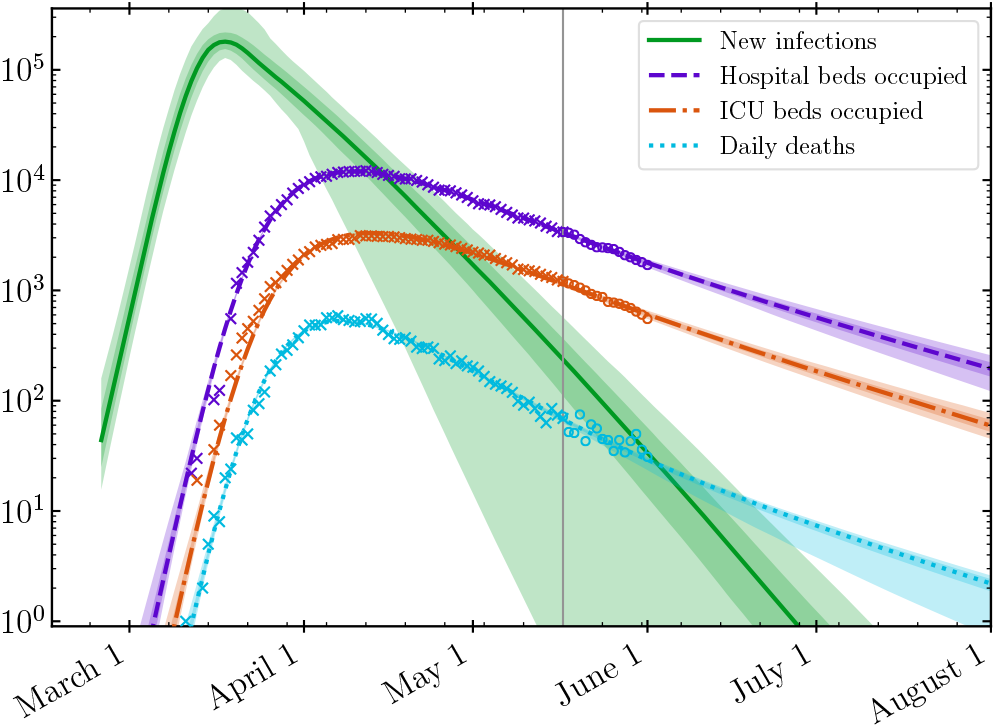
Model fit and prediction for hospital and ICU bed occupancy [55, 56] and deaths [57] in New York City using public data. Data from before the gray line on May 17, 2020 (crosses) are used for calibration. More recent data are plotted as circles. The model predictions shown here do not account for changes to mitigation after May 17, 2020.

### Spatial heterogeneity

In reality, the entire state of Illinois is not a single well-mixed system, even if such approximations are frequently made [2, 15, 16]. Illinois has a densely populated region in the northeast, and more sparsely populated regions further west and to the south. Our modeling identified that the status of the epidemic (and the projections for scenarios in which mitigation is relaxed) differ among the four super-regions. In particular, in the Southern and Central regions, the epidemic is declining more rapidly, due to a combination of social distancing and contact tracing using pre-existing resources. Modeling these regions separately, and including population transfer between them, is essential to guide the implementation of region-dependent mitigation strategies and to provide input to policy makers to prevent the resurgence of the epidemic.

### Relaxing mitigation

Our simulations suggest that it is too soon to lift all social distancing and mitigation restrictions, as the significant number of currently-infectious individuals would make a second wave inevitable. The dynamics of a second wave and the approach to herd immunity may be crucially sensitive to effects beyond a mean-field description. For example, we do not account for super-spreader events which have played a major role in the spread of SARS and MERS [10, 11, 13] and are likely also relevant for COVID-19 [58]. Similarly, we do not account for heterogeneities in the population structure. Such features may accelerate the onset of herd immunity which in turn decreases *R_t_*. These differences have the potential to decrease the severity of the second wave and may enable more efficient containment. We defer a treatment of these effects to future work.

### Additional limitations

We end with a brief discussion of additional limitations. First, the importance of spatial structure, heterogeneities in population susceptibility, and social network structure are well-appreciated in the literature [2, 12, 15]. We plan to investigate methods to model these effects and their impact on our results in future work. In addition, important sources of error in our existing model include the unknown impact of seasonal forcing, the discreteness of populations, and the effect of super-spreader events and behavioral response.

In our initial modeling [47], we used seasonal forcing with *A*_SF_ = 0.2 because similar effects had been documented for historical coronaviruses [19]. Because seasonal forcing varies slowly with time, it remains difficult to verify this assumption with the observables we consider. Nevertheless, our parameter inference does not exhibit strong evidence for this level of seasonal forcing; the role of climate in the early stages of epidemics is a question of active investigation [59, 60]. Furthermore, non-zero values for *A*_SF_ do not necessarily ascribe an equivalent seasonal modulation to the disease’s infectivity.

In addition, we wish to comment on the reported confidence intervals obtained from parameter inference for both stochastic and deterministic epidemic models [61], because these can depend on whether or not the model calibrations are performed for raw data or cumulative data. Generally speaking the estimated intervals are unrealistically small when models are fit to cumulative data [61], with stochastic models giving slightly larger intervals. Our calculations are calibrated to raw data not cumulative data, and so we do not expect the uncertainties in general to be estimated inappropriately. Regardless, any deterministic model of the epidemic trajectory will be inappropriate when few individuals are infected, in spite of apparent certainty in posteriors. Thus, in a regime where the deterministic model is not even appropriate as a first approximation, i.e., small populations, the estimate of uncertainties will not be a good indicator of the inapplicability of the deterministic model. Moreover, it is well-known in ecology that the discreteness of populations—the fact that individuals are quantized and that birth-death processes are discrete—leads to important qualitative phenomena ranging from population cycles [62] to noise-induced pattern formation [63, 64]. These effects are particularly important when numbers are small in the early and late stages of the epidemic because our mean-field model is incapable of representing the extinction state, i.e., when the number of infectious individuals drops below one. Once the epidemic is extinct in a particular region, it can only re-emerge due to migration events, e.g., a super-spreader event like when university students return to campus. In this regime, a prudent regional health department uses contact tracing to contain outbreaks. Such mechanisms are not represented in our modeling.

The third major limitation is that modeling an epidemic is very different from modeling a physical system, even one as complex as a weather pattern. The transmission of an infectious disease involves a collaboration between the virus and the host population: the host population alters its behavior in response to its awareness of epidemic progress, leading to policy steps that may increase or decrease transmission, and self-regulation of social distancing by susceptible and vulnerable populations. Thus, it is important to emphasize that predictions can easily be invalidated due to subsequent human actions that cannot be anticipated, and will be impossible to model precisely.

## VI. CONCLUSIONS

In this paper, we have presented a mathematical model and computational framework for recapitulating the COVID-19 epidemic. This model may be used to infer values for the parameters that drive and represent the progression of the disease. We use our calibrated model to provide robust short-term forecasts of the epidemic trajectory in different regions of the state and explore the effects that steps to relax social distancing measures may have, especially in the context of a second wave of the epidemic. The resulting highly-constrained and quantitative narrative of the epidemic is a useful tool to inform scenarios for sustainable monitoring and control of the epidemic.

## CODE AVAILABILITY

The model and calibration framework described above have been implemented in the open source Python 3 [65] package pydemic. The source code for pydemic is freely available online at https://github.com/uiuc-covid19-modeling/pydemic. This work made use of NumPy [66], SciPy [67], pandas [68], emcee [33], corner.py [69], and Matplotlib [70].

## Data Availability

The manuscript uses data provided by the Illinois Department of Public Health through a Data Use Agreement with Civis Analytics. The source
code for the model is freely available online at
https://github.com/uiuc-covid19-modeling/pydemic

## ACKNOWLEDGMENTS

We gratefully acknowledge discussions with David Ansell at Rush University Hospital, Mark Johnson at Carle Hospital, Katie Gostic and Sarah Cobey at University of Chicago, Jaline Gerardin at Northwestern University, Charles Gammie at the University of Illinois, and Josh Speagle at Harvard University. The calculations we have performed would have been impossible without the data kindly provided by the Illinois Department of Public Health through a Data Use Agreement with Civis Analytics. This work was supported by the University of Illinois System Office, the Office of the Vice-Chancellor for Research and Innovation, the Grainger College of Engineering, and the Department of Physics at the University of Illinois at Urbana-Champaign. Z.J.W. is supported in part by the United States Department of Energy Computational Science Graduate Fellowship, provided under Grant No. DE-FG02-97ER25308. This work made use of the Illinois Campus Cluster, a computing resource that is operated by the Illinois Campus Cluster Program (ICCP) in conjunction with the National Center for Supercomputing Applications (NCSA) and which is supported by funds from the University of Illinois at Urbana-Champaign. This research was partially done at, and used resources of the Center for Functional Nanomaterials, which is a U.S. DOE Office of Science Facility, at Brookhaven National Laboratory under Contract No. DE-SC0012704.

## APPENDIX

We detail the parameterization of our age-dependent severity model below. As noted in Section I A, we track the number of individuals in different age groups separately, and thus we specify the fiducial transition probabilities for each age group independently. We further define an overall scaling prefactor *p_x_* that governs the final transition probability *p_x,i_* as the product of the fiducial rate and the prefactor. Our fiducial transition rates *p_x,i_*/*p_x_* are drawn from Ref. [28,71-73].

In Table S1 we list the age-dependent values for the probabilities that: infected individuals experience symptoms *p_σ_*,*_i_*/*p_σ_*, symptomatic individuals are hospitalized *p_h_*,*_i_*/*p_h_*, hospitalized patients enter the ICU *p_c,i_*/*p_c_*, and ICU patients expire *p_d,i_/p_d_*. We also list the relative age distribution of the population in the United States, which we source from the UN [74]. As specified in Table I, we sample over the *p_c_* and *p_d_* scale factors.

**TABLE S1.** Table of age-specific parameters, including the age distribution *w_i_* and the various probabilities of transitioning between various states in our model. We present the unscaled distribution *p_x_*,*_i_/p_x_*, where *p_x_* denotes the overall scaling which is sampled during parameter inference (or whose value is otherwise set as described in the text).

| | [0, 10) | [10, 20) | [20, 30) | [30, 40) | [40, 50) | [50, 60) | [60, 70) | [70, 80) | [80, ∞) |
| --- | --- | --- | --- | --- | --- | --- | --- | --- | --- |
| $w_i$ | 0.120004 | 0.127891 | 0.139256 | 0.134948 | 0.121898 | 0.12725 | 0.116278 | 0.0727565 | 0.0397193 |
| $p_{\sigma,i}/p_\sigma$ | 0.057 | 0.054 | 0.294 | 0.668 | 0.614 | 0.83 | 0.99 | 0.995 | 0.999 |
| $p_{h,i}/p_h$ | 0.001 | 0.003 | 0.012 | 0.032 | 0.049 | 0.102 | 0.166 | 0.243 | 0.273 |
| $p_{c,i}/p_c$ | 0.05 | 0.05 | 0.05 | 0.05 | 0.063 | 0.122 | 0.274 | 0.432 | 0.709 |
| $p_{d,i}/p_d$ | 0.3 | 0.3 | 0.3 | 0.3 | 0.3 | 0.4 | 0.4 | 0.5 | 0.5 |

Because the data to which we calibrate does not constrain the symptomatic population, we cannot observe *p_h_* and so we fix it to one. We force our model to produce a given infection fatality ratio by setting *p_σ_* to

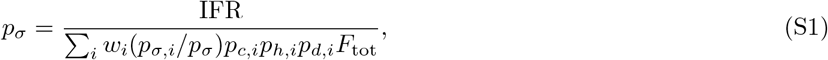

where the value for *p_σ,i_/p_σ_* on the right hand side of the equation is taken first from Table S1. This equation is valid if the initial infected population is distributed in proportion to *w_i_*.

In Table S2, we summarize the posterior probability distribution for all sample parameters enumerated in Table I.

Figure S1 shows joint posterior distributions for all pairs over the complete set of parameters of our model fitted to all-state data up to May 17, 2020 as shown in the bottom panel of Fig. 2.

**TABLE S2.** Table of inferred model parameters for disease severity and the time dependence of mitigation effects. Mitigation was modeled with a piecewise Hermite cubic interpolating polynomial as described in Section I A. Each column reports median parameter values from the MCMC algorithm for the different model fits along with the largest of the upper and lower one-sigma error bounds reported by the algorithm. In addition, we report the (derived) values and uncertainties of *R_t_* (as defined in Eq. 3) for each model as evaluated on May 1, 2020. The column headers specify the modeled region of Illinois and the inclusive end date of the calibration data.

|  | Illinois (5/17) | Illinois (4/20) | Northeastern (5/17) | North-Central (5/17) | Central (5/17) | Southern (5/17) |
| --- | --- | --- | --- | --- | --- | --- |
| $R_0$ | $2.2 \pm 0.26$ | $2.2 \pm 0.25$ | $2.3 \pm 0.28$ | $1.6 \pm 0.38$ | $1.6 \pm 0.4$ | $1.6 \pm 0.33$ |
| $t_s$ | Feb 15 ± 3.92 | Feb 14 ± 4.24 | Feb 16 ± 3.73 | Feb 16 ± 7.82 | Feb 18 ± 8.25 | Feb 17 ± 8.40 |
| $t_0$ | Mar 9 ± 4.13 | Mar 10 ± 4.13 | Mar 9 ± 4.04 | Mar 8 ± 5.71 | Mar 8 ± 6.08 | Mar 11 ± 7.62 |
| $t_1$ | Mar 20 ± 2.94 | Mar 21 ± 3.28 | Mar 20 ± 3.01 | Mar 24 ± 6.04 | Mar 25 ± 6.58 | Apr 4 ± 5.53 |
| $M(t_1)$ | $0.48 \pm 0.051$ | $0.47 \pm 0.048$ | $0.45 \pm 0.048$ | $0.64 \pm 0.12$ | $0.61 \pm 0.12$ | $0.59 \pm 0.099$ |
| $R_t(\text{May 1})$ | $0.94 \pm 0.01$ | $0.97 \pm 0.037$ | $0.94 \pm 0.011$ | $0.99 \pm 0.02$ | $0.95 \pm 0.023$ | $0.9 \pm 0.027$ |
| IFR | $0.007 \pm 0.0012$ | $0.0071 \pm 0.0012$ | $0.007 \pm 0.0012$ | $0.007 \pm 0.0012$ | $0.007 \pm 0.0012$ | $0.007 \pm 0.0012$ |
| $\tau_h$ | $6.3 \pm 2$ | $6.2 \pm 2$ | $6.2 \pm 2$ | $6.1 \pm 1.8$ | $6.2 \pm 1.8$ | $6.9 \pm 1.8$ |
| $\sigma_h$ | $2.3 \pm 1.4$ | $2.2 \pm 1.6$ | $2.4 \pm 1.7$ | $4.2 \pm 1.9$ | $4.2 \pm 1.9$ | $4.6 \pm 1.7$ |
| $\tau_{\text{disch}}$ | $7 \pm 1.9$ | $6.9 \pm 1.9$ | $7.1 \pm 1.9$ | $6.2 \pm 1.7$ | $6.6 \pm 1.8$ | $6.8 \pm 1.9$ |
| $\sigma_{\text{disch}}$ | $6.1 \pm 1.8$ | $5.7 \pm 2$ | $6.4 \pm 1.7$ | $3.5 \pm 1.7$ | $4.6 \pm 1.9$ | $5 \pm 1.8$ |
| $p_c$ | $0.61 \pm 0.16$ | $0.67 \pm 0.18$ | $0.62 \pm 0.17$ | $0.47 \pm 0.14$ | $0.32 \pm 0.097$ | $0.51 \pm 0.14$ |
| $\tau_c$ | $1.4 \pm 0.97$ | $1.7 \pm 1.3$ | $1.3 \pm 0.93$ | $3 \pm 1.6$ | $2.2 \pm 1.5$ | $2 \pm 1.3$ |
| $\sigma_c$ | $2.2 \pm 1.8$ | $2 \pm 1.8$ | $2.3 \pm 1.9$ | $2.5 \pm 1.6$ | $2.5 \pm 1.7$ | $2.7 \pm 1.7$ |
| $p_d$ | $1.4 \pm 0.15$ | $1.3 \pm 0.16$ | $1.4 \pm 0.16$ | $1.6 \pm 0.3$ | $1.9 \pm 0.25$ | $1.9 \pm 0.26$ |
| $\tau_d$ | $8.8 \pm 0.96$ | $7.7 \pm 1$ | $8.8 \pm 0.98$ | $13 \pm 2.1$ | $8.9 \pm 1.6$ | $11 \pm 1.4$ |
| $\sigma_d$ | $4.6 \pm 1.5$ | $3.7 \pm 1.5$ | $4.7 \pm 1.6$ | $9.2 \pm 2.4$ | $7 \pm 2.7$ | $6.6 \pm 2.2$ |
| $\tau_{\text{rec}}$ | $11 \pm 2.3$ | $12 \pm 2.5$ | $11 \pm 2.3$ | $12 \pm 2.8$ | $12 \pm 2.8$ | $12 \pm 2.8$ |
| $\sigma_{\text{rec}}$ | $6.8 \pm 2.7$ | $8.7 \pm 2.9$ | $6.9 \pm 2.7$ | $10 \pm 2.8$ | $10 \pm 2.8$ | $9.9 \pm 2.9$ |
| $F_{\text{tot}}$ | $1.4 \pm 0.033$ | $1.3 \pm 0.06$ | $1.4 \pm 0.034$ | $1.2 \pm 0.14$ | $1.6 \pm 0.3$ | $2.1 \pm 0.28$ |
| $\tau_{\text{tot}}$ | $2.9 \pm 0.46$ | $2 \pm 0.53$ | $2.8 \pm 0.47$ | $2.2 \pm 0.88$ | $2.3 \pm 0.91$ | $2.8 \pm 0.88$ |
| $\sigma_{\text{tot}}$ | $2.3 \pm 0.88$ | $1.9 \pm 0.84$ | $2.2 \pm 0.86$ | $2.1 \pm 0.91$ | $2 \pm 0.92$ | $1.9 \pm 0.91$ |
| $A_{\text{SF}}$ | $0.15 \pm 0.03$ | $0.06 \pm 0.071$ | $0.11 \pm 0.035$ | $0.13 \pm 0.065$ | $0.12 \pm 0.07$ | $0.13 \pm 0.073$ |

**TABLE S3.**
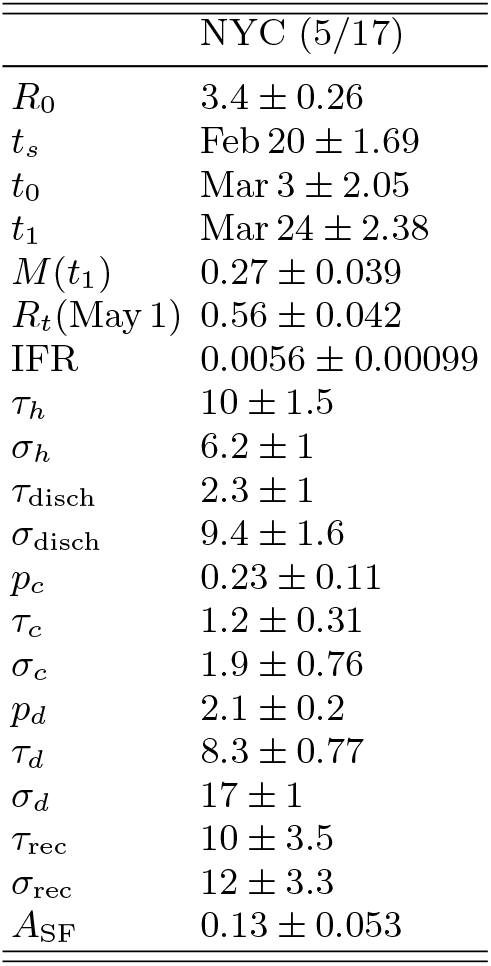
Same as Table S2, but for fits for New York City.

|  | NYC (5/17) |
| --- | --- |
| $R_0$ | $3.4 \pm 0.26$ |
| $t_s$ | Feb 20 ± 1.69 |
| $t_0$ | Mar 3 ± 2.05 |
| $t_1$ | Mar 24 ± 2.38 |
| $M(t_1)$ | $0.27 \pm 0.039$ |
| $R_t(\text{May 1})$ | $0.56 \pm 0.042$ |
| IFR | $0.0056 \pm 0.00099$ |
| $\tau_h$ | $10 \pm 1.5$ |
| $\sigma_h$ | $6.2 \pm 1$ |
| $\tau_{\text{disch}}$ | $2.3 \pm 1$ |
| $\sigma_{\text{disch}}$ | $9.4 \pm 1.6$ |
| $p_c$ | $0.23 \pm 0.11$ |
| $\tau_c$ | $1.2 \pm 0.31$ |
| $\sigma_c$ | $1.9 \pm 0.76$ |
| $p_d$ | $2.1 \pm 0.2$ |
| $\tau_d$ | $8.3 \pm 0.77$ |
| $\sigma_d$ | $17 \pm 1$ |
| $\tau_{\text{rec}}$ | $10 \pm 3.5$ |
| $\sigma_{\text{rec}}$ | $12 \pm 3.3$ |
| $A_{\text{SF}}$ | $0.13 \pm 0.053$ |

**FIG. S1.**
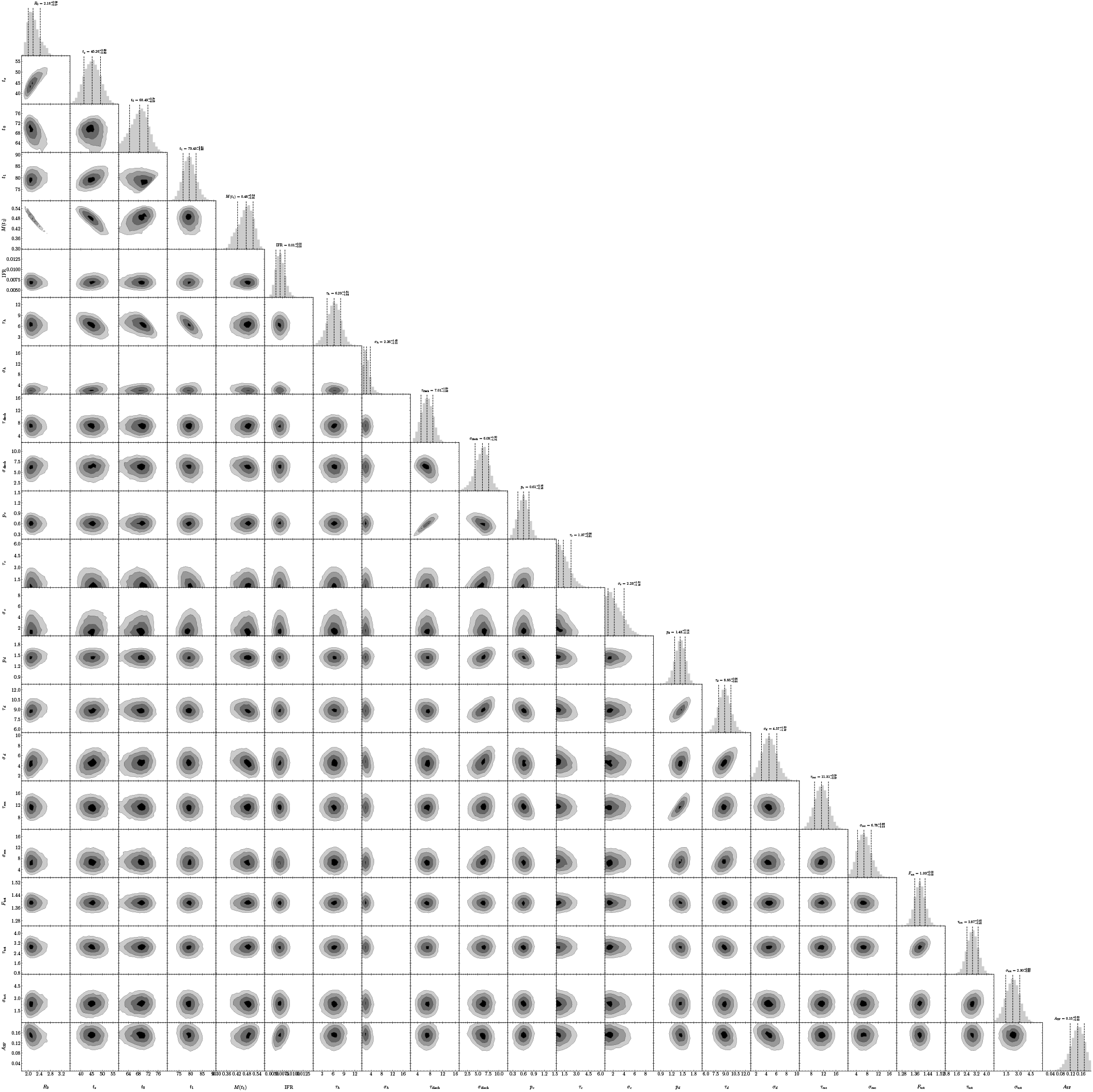
Joint posterior distributions of pairs of the complete set of parameters of our model fitted to the all-state data up to May 17, 2020. The correlations between some pairs of fitted parameters such as, e.g., between *R*_0_ and that start date of the epidemic are reflected in the ellipsoidal shape of the posteriors. This is an expanded version of Fig. 5

